# Insight and symptoms severity in schizophrenia explained by the flexibility of brain dynamics and pharmacological treatment

**DOI:** 10.64898/2026.04.30.26352132

**Authors:** Daniela Janeva, Martin Breyton, Jean Philippe Ranjeva, Raphael Richieri, Laurent Boyer, Maxime Guye, Christophe Lancon, Olivier Blin, Viktor Jirsa, Spase Petkoski, Romain Guilhaumou

## Abstract

Schizophrenia’s substantial heterogeneity poses a major challenge for understanding its neurobiological mechanisms and predicting treatment response. Moving toward precision psychiatry, we identified clinically meaningful subtypes and characterised their neural and pharmacological profiles. Clustering of multidimensional clinical feature space revealed two distinct patient subtypes, primarily differentiated by degree of illness insight. In parallel, three symptom-severity groups defined by positive and negative psychopathology dimensions provided a complementary stratification framework. Resting-state fMRI analyses revealed that higher-insight patients exhibited greater dynamic reconfiguration of regional functional connectivity, emerging as the primary neuroimaging feature differentiating subtypes. Multivariate classification and feature importance analysis confirmed the discriminative value of neuroimaging metrics. Across both subtyping approaches, regional flexibility was spatially associated with cortical receptor density maps in a subtype-specific manner, particularly for D_**2**_ and 5-HT_**2*A***_ when accounting for estimated antipsychotic receptor occupancies. Additionally, pharmacological–clinical associations were stronger and more spatially widespread in specific subtypes, indicating subtype-dependent pharmacodynamic relationships. Furthermore, structural equation modelling demonstrated that neuroimaging measures mediate receptor pharmacology’s influence on clinical outcomes. These findings together show that integrating clinical, neuroimaging, and pharmacological data can uncover biologically grounded schizophrenia subtypes, identify functional biomarkers, and inform personalised therapeutic strategies.

## Introduction

Schizophrenia is an incapacitating psychiatric disorder characterised by a high degree of clinical and biological heterogeneity [1]. Its pathophysiological complexity challenges diagnostic precision and complicates the selection of appropriate pharmacotherapy and the prediction of treatment response [2]. Current pharmacological treatments are effective in only thirty to fifty percent of patients with schizophrenia, with a substantial proportion experiencing no therapeutic benefits or having adverse effects [3]. Together, these limitations underscore the need for personalised treatment strategies through the identification of more homogeneous phenotypic targets [4].

Schizophrenia is symptomatically characterised by a heterogeneous constellation of *positive* (e.g., hallucinations, delusions) and *negative* symptoms (e.g., affective flattening, anhedonia, social withdrawal). To quantify these dimensions, the *Positive and Negative Syndrome Scale* (PANSS) [5], provides standardised measures across positive, negative, and general psychopathology domains. In addition, a particularly relevant and underexplored dimension of schizophrenia heterogeneity is illness *insight*, the degree to which patients are aware of their own psychopathology, treatment need, and symptom attribution. Impaired insight is one of the most prevalent features of schizophrenia, affecting the majority of patients, and is independently associated with poorer medication adherence, worse functional outcomes, and greater symptom severity [6, 7]. The *Scale to Assess Unawareness of Mental Disorder* (SUMD) provides a standardised, multidomain evaluation of illness insight and is the primary instrument used to characterise patient awareness [8].

A critical first step toward personalised treatment is the robust definition of population subtypes that reflects both clinical manifestations and underlying neurobiological mechanisms [9]. Prior attempts to define schizophrenia subgroups are mostly based on similarities in cognitive and symptomatic profiles [10]. Although DSM-5 no longer includes subtype categorisation, symptom profiles are hypothesised to reflect underlying neurobiological mechanisms and should therefore remain central to subtyping efforts. Three major neurobiological systems—the dopaminergic, serotonergic, and glutamatergic are consistently implicated in schizophrenia’s pathophysiology and clinical manifestations [11–13]. Based on neuropsychopharmacological network models, dopaminergic dysregulation, specifically mesolimbic hyperactivity, was examined in relation to positive psychotic symptoms, while serotonergic dysfunction, through interactions with dopaminergic and glutamatergic pathways, contributes to cortical and cognitive impairments [14]. Among these systems, dopaminergic and serotonergic mechanisms have received the greatest translational attention, given their direct relevance to antipsychotic pharmacology. The dopamine D_2_ receptor remains central to antipsychotic efficacy, while other dopamine receptor subtypes (D_1_, D_3_, D_4_) contribute to cognitive modulation, reward processing, and interindividual variability in treatment response [11, 12]. Serotonergic receptor subtypes, notably 5-HT_1A_, 5-HT_2A_, 5-HT_2C_, and 5-HT_6_, are key modulators of mood, cognition, and treatment tolerability, and support the pharmacodynamic rationale of second-generation antipsychotics, which combine serotonin and dopamine antagonism to improve efficacy while limiting side effects [11, 15].

Blood oxygen level–dependent (BOLD) functional magnetic resonance imaging (fMRI) provides an indirect measure of neural activity through neurovascular coupling, reflecting changes in local deoxyhemoglobin concentration associated with synaptic and metabolic demand [16]. BOLD is widely used to characterise large-scale brain network dynamics in schizophrenia [17–19], with neuromodulatory systems such as dopaminergic and serotonergic projections, shaping the gain, stability and influencing the macroscopic dynamics captured by resting-state fMRI (rs-fMRI) [20, 21]. This distinction is particularly critical in medicated schizophrenia cohorts. Antipsychotics exert their therapeutic effects primarily through dopamine D_2_ receptor blockade and serotonergic modulation, altering synaptic transmission and large-scale network organisation [11, 12]. Consequently, resting-state functional patterns acquired under ongoing treatment likely reflect a superposition of intrinsic pathophysiology and pharmacologically induced neuromodulation. Disentangling these contributions remains a major challenge for neuroimaging studies of schizophrenia and limits the interpretability of subtype-specific neural signatures.

Traditionally, subtypes have been described according to positive and negative symptom dimensions or cognitive performance [22]. More recent approaches have sought to integrate multimodal data—combining clinical, neuroimaging, and genetic dimensions—to reveal neurobiologically distinct patient subgroups [23–25]. Despite these advances, a comprehensive framework integrating clinical phenotypes with their neural correlates and pharmacological targets remains lacking [17, 18]. Here, we present an integrative framework structured around three complementary objectives: characterising neuroimaging and pharmacological profiles in (I) data-driven clinical subtypes, (II) symptom-severity groups defined by PANSS profiles, and (III) a multimodal integration examining inter-subtype differences across data modalities. Through this framework, we aim to identify homogeneous phenotypic profiles that reflect underlying pathophysiological differences and address a central question: how do pharmacological effects manifest in neural dynamics, and can these differences refine our understanding of schizophrenia heterogeneity and inform personalised therapeutic strategies?

## Results

To identify neural and pharmacological schizophrenia profiles and their causal influence on symptom expression, we performed intergroup analysis of clinical schizophrenia subtypes by integrating multimodal data of the same cohort. Analyses were conducted on data from 92 stable schizophrenic patients aged 18–50, among them, 87 had neuroimaging data, 85 had pharmacological data, and 83 had complete multimodal datasets (Table 1). All patients were considered clinically stable for 6-months before inclusion and were treated by antipsychotics. Thus, dissociating disease-related alterations from treatment-associated neuromodulation is critical for interpreting functional differences. To quantify treatment-related effects, we estimated receptor occupancy using a pharmacokinetic–pharmacodynamic (PK–PD) approach, enabling subject-specific inference of dopaminergic and related neuromodulatory engagement. Resting-state fMRI data were minimally preprocessed, and functional features were extracted using the Schaefer 7-network, 100-region cortical parcellation incorporating 18 subcortical regions. From these data, we derived 819 functional features capturing whole-brain, network-specific, and regional measures of functional connectivity integration and segregation, mean and variance of functional connectivity dynamics, metastability, and regional signal complexity measures: ALFF, fALFF, and zero-crossing rate. The observed functional differences are expected to reflect a combination of disease-related alterations and treatment-induced neuromodulatory effects. These features formed the basis for all subsequent analyses, allowing us to investigate multimodal interactions addressing clinical heterogeneity in schizophrenia.

**Table 1.** Descriptive statistics of the study population. Values are mean ± SD unless otherwise specified. n = 92.

| Variable | Overall | Variable | Overall |
| --- | --- | --- | --- |
| *Demographics* |  | *Insight (SUMD items)* |  |
| Age (years) | 30.1 ± 7.5 | Item 1 | 1.6 ± 0.8 |
| Illness duration (years) | 8.8 ± 5.9 | Item 2 | 1.7 ± 0.9 |
| Onset to treatment (years) | 1.1 ± 2.6 | Item 3 | 1.5 ± 0.8 |
| *Comorbidities and risk factors (No/Yes)* | | Item 4 | 0.9 ± 1.0 |
| Alcoholism | 76 / 16 | Item 5 | 1.4 ± 1.1 |
| Drug addiction | 81 / 11 | Item 6 | 1.6 ± 1.0 |
| Cannabis use | 42 / 50 | Item 7 | 1.6 ± 0.9 |
| Smoking | 34 / 58 | Item 8 | 1.6 ± 0.9 |
| Suicide attempt | 67 / 23 | Item 9 | 1.6 ± 0.9 |
| *Symptom severity* |  | *Medication, n (%)* |  |
| PANSS Positive | 13.9 ± 5.9 | Aripiprazole | 22 (22.8) |
| PANSS Negative | 24.4 ± 8.1 | Clozapine | 15 (16.3) |
| PANSS General | 36.0 ± 10.4 | Olanzapine | 13 (14.1) |
| Calgary Depression Scale | 1.4 ± 3.1 | Risperidone | 10 (12.0) |
| CGI | 4.1 ± 1.5 | Amisulpride | 8 (8.7) |
| GAF | 59.8 ± 16.9 | Quetiapine | 7 (8.7) |
|  |  | Risperidone consta | 7 (7.6) |
|  |  | Haloperidol decanoate | 3 (3.3) |

### Rs-fMRI features and receptor occupancy profiles distinguish symptom-defined schizophrenia subtypes

We took two approaches to define clinically meaningful subtypes (Fig. 1a). First, two robust clinical data-driven subtypes were identified by performing unsupervised learning on pre-processed clinical data represented in Table 1. The *k-means* algorithm was applied to a 43-dimensional clinical dataset, and multiple clustering validity indices were evaluated to determine the optimal number of clusters. This approach revealed two well-separated subtypes, primarily distinguished along the first principal component, accounting for 17.5% of the total variance (Fig. 1b, left). As this component was predominantly driven by SUMD scale items (Fig. 1c, left), the subtypes were defined as High Insight and Low Insight. Secondly, we defined hypothesis-driven subtypes according to the PANSS scale by selecting specific thresholds. We defined three groups: Low Positive Low Negative (LPLN), Low Positive High Negative (LPHN), and High Positive High Negative (HPHN) (Fig. 1b, right). 92.31% of patients in the LPLN group, 66.67% in the LPHN group, and 39.29% in the HPHN group were classified as High Insight (Fig. 1c, right).

**Fig. 1.**
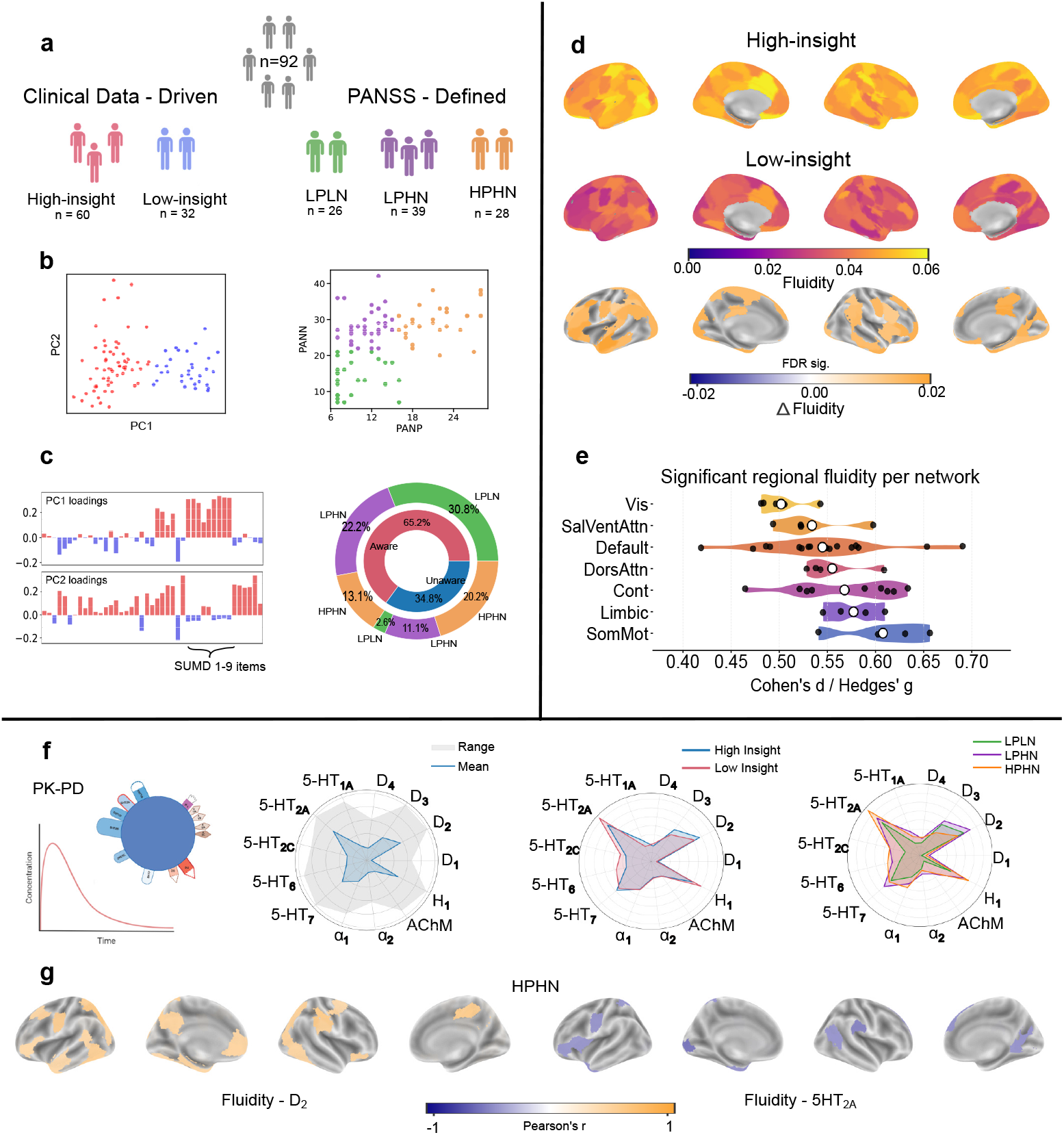
Clinical subtypes, functional connectivity fluidity, and pharmacological profiles in schizophrenia. **(a)** Two Clinical data-driven subtypes, High Insight and Low Insight (left) and three PANSS-defined groups derived from the same cohort. **(b)** Clinical data-driven clusters represented in lower-dimensional space, separated by the PC1 (left) and PANSS groups represented by PANP and PANN dimension on the x and y axis, respectively. **(c)** (left) PC components defining the lower-dimensional space, PC1 governed by the SUMD scale, and overlap between the subtypes (right). **(d)** Regional functional connectivity, variability is consistently higher in High Insight than Low Insight patients. Group differences were assessed using the Mann–Whitney *U* test with FDR correction. **(e)** Effect sizes (Cohen’s *d* / Hedges’ *g*) confirmed significant differences across all functional networks, with the largest representation in the Default Mode Network and the highest mean values in the Somatomotor Network. **(f)** Mean PK-PD estimation of receptor occupancy in the whole dataset, and in subtypes, respectively. Across all subgroups, 5-HT_2*A*_ receptors showed the highest mean occupancy. **(g)** Subtype-specific spatial patterns of significant fMRI-receptor correlations within the HPHN subtype.

From the rs-fMRI dataset, we computed functional neuroimaging features (Section 7). Among them, regional fluidity (Fig. 6) emerged as the most discriminative feature between the High and Low Insight subtypes, assessed by two-sided Mann–Whitney U tests with False Discovery Rate (FDR) correction (*q* < 0.05). We chose a non-parametric approach due to the relatively small sample sizes within each clinical subtype, which may violate assumptions of normality required for parametric tests. When focusing on cortical regional fluidity, 48 out of 100 regions survived FDR correction, all of them showing higher fluidity for the High Insight subtype (Fig. 1b). Spatial mapping of regional fluidity further demonstrated systematically higher values in the High Insight subgroup relative to the Low Insight subgroup (mean difference = 0.016, SD = 0.002, *q*_min_ = 0.010) (Fig. 1d), indicating reduced functional flexibility in patients with impaired clinical insight. All significant regional effects were positive, supporting a consistent directionality of lower fluidity in the Low Insight subgroup. Effect sizes quantified using Cohen’s d and Hedges’ g were distributed across all functional networks, with the largest number of significant regions arising from the Default Mode Network (*mean* |*d*| = 0.544) and higher average effects observed in somatomotor regions (*mean* |*d*| = 0.607) (Fig. 1e).

In contrast, multigroup comparisons across PANSS-defined subtypes using Kruskal–Wallis tests on the full neuroimaging features dataset did not yield significance after FDR correction (all *q* > 0.05), and post-hoc Mann-Whitney U test did not converge to any feature set consistently differentiating the three hypothesis-driven subtypes. Focusing on the cortical regional fluidity, significant differences are observed in a few regions between the LPLN and LPHN subtypes (Kruskal-Wallis *p* > 0.05, post-hoc Man-Whitney FDR) as shown in the Appendix A (Fig. A1). This indicates that neural profiles do not differ significantly between PANSS-defined groups, likely reflecting overlapping patterns across symptom severity. However, the limited sample size, particularly when subdivided into three groups, may have reduced statistical power to detect subtle between-group differences.

We estimated receptor occupancy for each individual based on the prescribed dosage and medication profile. Using a pharmacokinetic–pharmacodynamic framework that accounts for free plasma concentration and receptor binding affinities, receptor occupancy was computed as a fractional measure representing the global brain-level engagement of each target receptor (Section 7). Thirteen receptors were included, covering dopaminergic and serotonergic pathways involved in schizophrenia pathophysiology, as well as receptors commonly associated with treatment-related side effects. The mean receptor occupancies for the whole population differ from subtype-specific means (Fig. 1f). Most notable correlations are represented in the extreme symptomatological group (Fig. 1g). Within the HPHN subtype, significant positive associations were observed between *D*_2_ receptor density and fluidity, whereas 5 − *HT*_2*A*_ receptor density showed significant negative correlations with fluidity. Regarding the ALFF feature, both groups exhibit positive correlations with dopaminergic and serotonergic receptors. More information on the neuroimaging-pharmacological relations can be found in Fig. A4 -Fig. A6. These results serve as a proxy for subtype-specific systematic modulation of intrinsic brain dynamics.

### Functional neuroimaging features robustly discriminate clinical subtypes

Focusing on the ability of rs-fMRI to differentiate the clinical subtypes, we performed a classification task on the 819-dimensional functional feature space. By assigning the cluster labels as ground truth, we implemented a nested cross-validation Support Vector Machine (SVM) classification (Section 7) preceded by normalisation and feature selection. Feature importance was further assessed using SHAP values and permutation scores. Averaging the classification performance scores across the outer loops offers a more reliable assessment of model performance on small datasets (Table 2). Applying this pipeline to the complete feature set, the primary classifier applied to the whole dataset (SVM 1) achieved robust performance, with an accuracy of 0.76 ± 0.07, balanced accuracy of 0.72 ± 0.09, macro-F1 score of 0.72 ± 0.09, and AUC of 0.78 ± 0.13. Subsequently, when the classifier was restricted to the most statistically significant features surviving Mann-Whitney *U* test with FDR correction (*q* < 0.05), which were predominantly regional fluidity measures (Fig. 2b) the SUMD model (SVM 2) remained above chance level, achieving 0.67 ± 0.12 accuracy and 0.69 ± 0.14 balanced accuracy. This feature selection step reduced the input dimensionality substantially while preserving the core discriminative signal, demonstrating that the compact subset of fluidity features specifically retains substantial discriminatory information. The consistency of classification performance between the full and reduced feature sets further suggests that the statistically significant fluidity regions capture the most relevant neural variance for distinguishing High Insight from Low Insight subtypes. Applying the same classification framework to multiclass differentiation of PANSS-defined subgroups resulted in lower performance (SVM 3, accuracy 0.58±0.09, balanced accuracy 0.57±0.10, normalised), suggesting a more overlapping organisation of these categories in functional neuroimaging space. The comparison between the classification performance of the first two models outlines the power of regional fluidity, framing it as a potential biomarker.

**Table 2.** Nested cross-validation performance of the SVM classifiers for SUMD and PANSS classifications. Values are mean ± standard deviation across outer folds.

| Model | Accuracy | Balanced Accuracy | F1 (macro) | AUC |
| --- | --- | --- | --- | --- |
| SVM 1 (SUMD) | 0.76 ± 0.07 | 0.72 ± 0.09 | 0.72 ± 0.09 | 0.78 ± 0.13 |
| SVM 2 (SUMD) | 0.67 ± 0.12 | 0.69 ± 0.14 | 0.66 ± 0.13 | 0.72 ± 0.09 |
| SVM 3 (PANSS) | 0.58* ± 0.09* | 0.57* ± 0.10* | 0.55* ± 0.09* | — |

\* Normalized with respect to chance level.

**Fig. 2.**
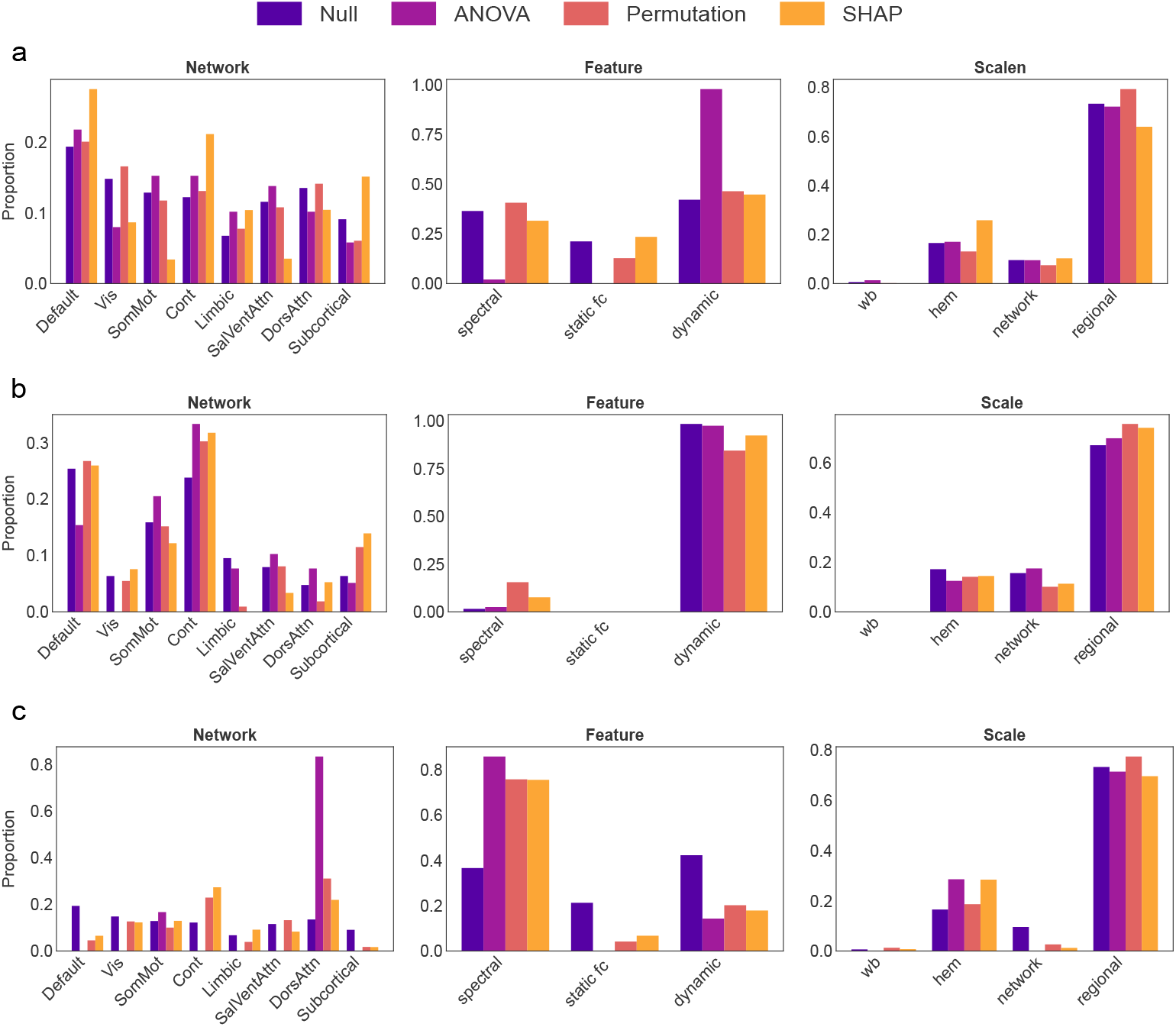
Feature importance distributions across classification frameworks and importance measures. Normalized proportion of feature categories across functional network (left), feature type (middle), and spatial scope (right) for three support vector machine (SVM) classifiers: (a) SVM1, binary classification of High versus Low Insight using all neuroimaging features; (b) SVM2, binary classification of High versus Low Insight using Mann–Whitney U preselected features; and (c) SVM3, multiclass classification of PANSS-defined subgroups. Each panel shows four normalised distributions: null (full feature space), ANOVA (features consistently selected across cross-validation folds), permutation importance (normalised summed accuracy-decrease scores), and SHAP (normalised mean absolute SHAP values). Divergence between ANOVA and model-based measures (permutation, SHAP) reflects disagreement between univariate filtering and multivariate predictive contribution. SVM1 emphasises Default Mode Network dynamic regional features; SVM2 highlights dynamic Control Network features; SVM3 is characterised by Dorsal Attention Network and spectral features. Occasional divergence between permutation and SHAP values reflects differences between marginal and interaction-dependent feature contributions.

To characterise the selectivity of each importance measure, we compared the distribution of top features across three organisational axes — functional network, feature type (spectral, static FC, dynamic FC,), and spatial scope (whole-brain, network, hemisphere, regional) — against a null baseline reflecting the composition of the full feature space. For each axis, we derived four distributions (null, ANOVA, permutation importance, SHAP) expressed as proportions summing to 1, allowing direct comparison of feature enrichment relative to base rates. This approach distinguished features that are merely frequent in the data (null), those passing a univariate relevance threshold (ANOVA), and those contributing to multivariate predictive behaviour (permutation importance and SHAP). Divergence between the ANOVA and importance-based distributions revealed cases where univariate filtering and model-based importance disagree on which feature categories drive predictions. The SVM1 binary classification of all neuroimaging features into High Insight and Low Insight groups revealed that features related to the Default Mode Network were most influential, particularly dynamic and regional measures (Fig. 2a). In contrast, SVM2, which employed features preselected using the Mann-Whitney U test, primarily highlighted dynamic features associated with the Control Network as most informative for the same classification task (Fig. 2b). The SVM3 multiclass classification of PANSS-defined subgroups emphasised the importance of the Dorsal Attention Network. Unlike the insight-based classifications, spectral features emerged as the most discriminative in this framework (Fig. 2a). Although the majority of selected features were regional, their relative importance increased after normalisation by proportion. Also, it is important to distinguish between permutation importance and SHAP values. Permutation scores quantify the independent contribution of individual features, whereas SHAP scores capture feature importance in the context of interactions with other features and are not always consistent with one another. The insight is evidently represented by the dynamic features, but given the prominence of spectral features in the PANSS-based classification, we further examined inter-subtype cortical differences (Appendix A; Fig. A2, Fig. A3), illustrating between-group cortical differences in the clinical data-driven and PANSS-defined subtypes, respectively. Overall, these results demonstrate that functional neuroimaging features reliably discriminate clinically meaningful subtypes. Classification performance remained above chance even when restricted to the most informative features, supporting their potential as candidate neural biomarkers underlying clinical heterogeneity.

### Subtype-specific cross-domain associations and model-based integration

Correlation analysis (Pearson’s correlation coefficient *r*, two-sided tests with FDR correction) revealed a pronounced heterogeneity in the association between receptors’ occupancies and clinical variables across symptom-defined subtypes. Weak and sparse correlations were observed in the full cohort, most notably a negative association between D_2_/D_3_ occupancy and SUMD1-3 (Fig. 3). The LPLN subtype showed limited pharmacological-clinical associations, with only a small number of significant correlations, predominantly involving D_1_ and SUMD2 (negative) and D_3_ with SUMD6 (positive), suggesting a relatively attenuated receptor-symptom coupling in this low-burden subgroup. In contrast, the LPHN subtype exhibited selective correlations between serotonergic receptor occupancy and positive symptom measures, particularly between 5-HT_1*A*_ and PANP, with an otherwise sparse correlation landscape suggesting that receptor occupancy explains only a limited portion of the clinical variance in this subtype. The Low Insight subtype displayed a distinct pattern characterised by associations involving both dopaminergic and serotonergic receptors and cognitive-affective symptoms, with D_2_ showing a positive correlation with SUMD6. Notably, the HPHN subtype showed the highest density and magnitude of significant correlations (|*r*| ≈ 0.4–0.6, FDR-adjusted *p* < 0.05), encompassing multiple receptor systems and a broad range of clinical measures. Within this subtype, D_1_ showed opposing associations with SUMD5, D3 was strongly linked to many of the SUMD components. These findings indicate that pharmacological-clinical relationships are largely obscured at the group level but emerge in a subtype-dependent manner, underscoring the importance of stratification for resolving heterogeneous drug–symptom coupling in schizophrenia. The full correlation heatmaps are shown in Appendix A (Fig. A7).

**Fig. 3.**
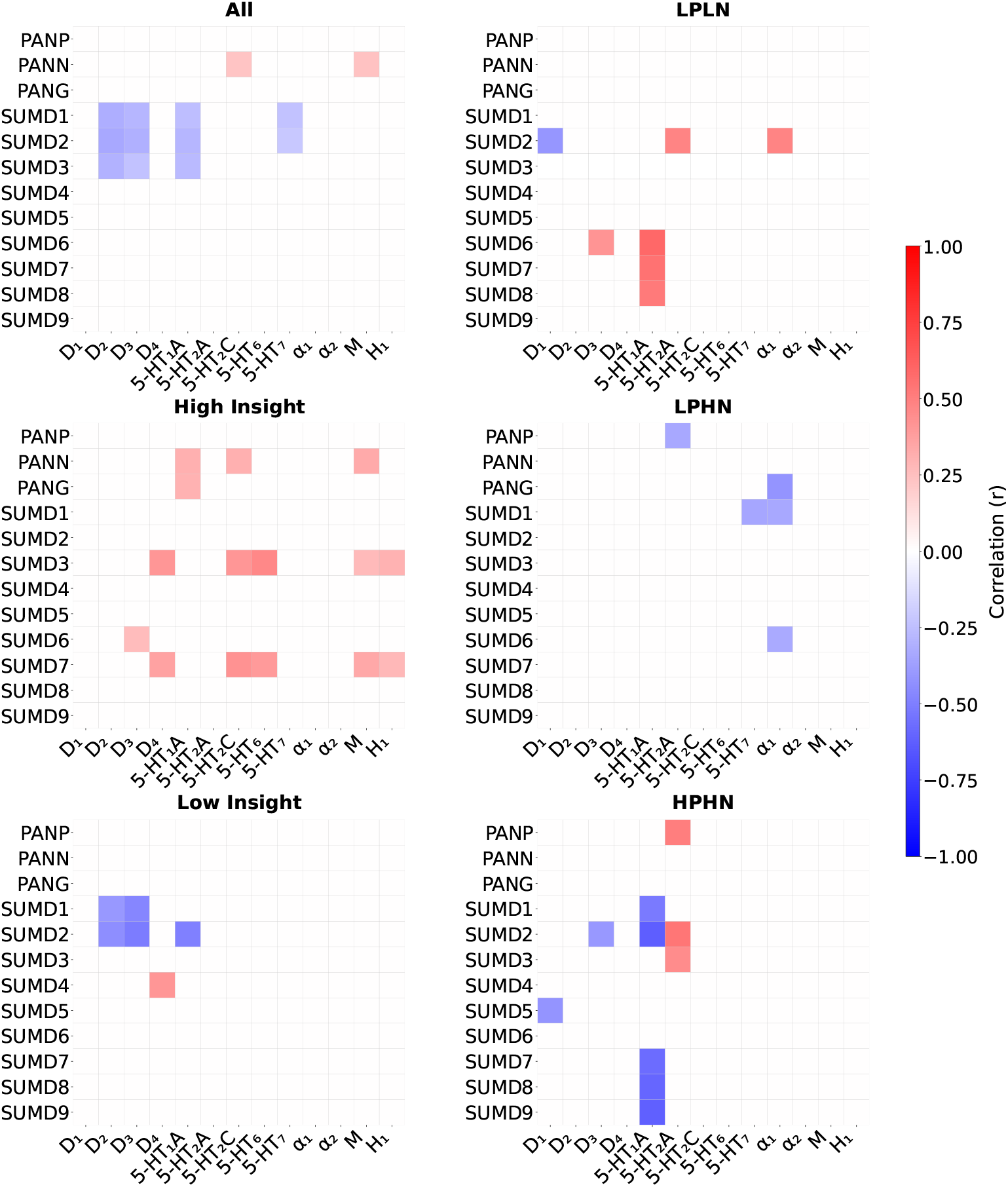
Subtype-specific correlations between receptor occupancy and clinical variables. Heatmaps show Pearson correlation coefficients between estimated receptor occupancy and clinical measures for the full cohort (All) and for each symptom-defined subtype (High/ Low Insight, LPLN, LPHN, HPHN). Only correlations surviving false discovery rate (FDR) correction across receptors and clinical variables are displayed. Colour intensity indicates the strength and direction of the association, with red denoting positive and blue denoting negative correlations. Subtype stratification reveals distinct and non-overlapping correlation patterns that are largely obscured in the full cohort, with the highest density and magnitude of significant associations observed in the HPHN subtype.

**Fig. 4.**
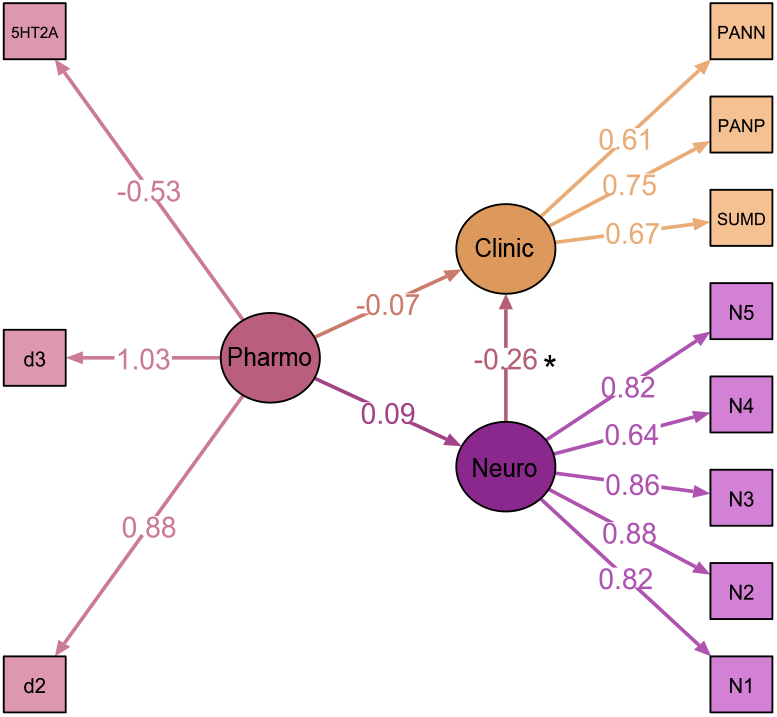
Structural equation model integrating pharmacological, neuroimaging, and clinical domains. Schematic representation of the hierarchical SEM relating latent pharmacological, neuroimaging, and clinical constructs. The latent Pharmacology factor is defined by estimated receptor occupancy measures, the latent Neuroimaging factor by rs-fMRI–derived features, and the latent Clinical factor by symptom severity measures. Directed paths represent hypothesised dependencies between domains, with pharmacological effects modelled as influencing neuroimaging dynamics and clinical outcomes, and neuroimaging dynamics additionally conveying a pathway to clinical expression. Path coefficients shown are standardised estimates obtained using robust maximum likelihood estimation. Model fit indices indicate excellent agreement with the data (CFI = 0.985; RMSEA = 0.047). Marginally significant directional path from Latent Neuro to Latent Clinical (*p* = 0.043).

To formally assess cross-domain dependencies, incorporating neuroimaging as a mediator between pharmacological and clinical dimensions, we next estimated a hierarchical structural equation model (SEM). In this triangular model, a latent Pharmacology factor is defined by the D_2_, D_3_ and 5HT_2*A*_ receptor occupancies and was specified to predict both a latent Neuroimaging factor and a latent Clinical factor. The third link conveyed a directional pathway from the Neuroimaging factor to the Clinical one. The latent Neuroimaging included five features: mean and variance of functional connectivity dynamics (FCD) across both intra- and inter-regional scales. Specifically, inter-regional mean fluidity was extracted from the left Default Mode Network prefrontal cortex (parcels 3 and 4), the right Control Network lateral PFC (parcel 2), and intra-regional mean fluidity from the Somatomotor network. Inter-regional variance of fluidity was measured in the right Dorsal Attention Network precentral/ventral region (parcel 1). These neuroimaging features emerged as most discriminating. The latent Clinical variable was defined by the sum of Positive and Negative items and the sum of SUMD items. This formulation enabled an explicit test of whether pharmacological variation influences clinical expression directly or indirectly through neurofunctional dynamics.

On the full dataset, SEM demonstrated excellent fit to the data (CFI = 0.985; RMSEA = 0.047). Functional connectivity dynamics emerged as a significant predictor of clinical severity (*β* = −0.26, *P* = 0.041), where the standardised coefficient indicates that an increase of one standard deviation in neural dynamics was associated with a 0.26 standard deviation reduction in clinical severity, reflecting an inverse relationship, concurrently greater or more intact neural activity and connectivity corresponded to lower symptom burden. In contrast, the pharmacological latent factor showed no direct association with clinical outcomes. These results suggest that inter-individual variability in global functional brain dynamics, reflecting a combination of underlying disease pathology and pharmacological engagement, mediates the relationship between neurochemical systems and symptom presentation, rather than pharmacological receptor modulation alone. Notably, the absence of significant direct paths from the pharmacological latent factor to neuroimaging and clinical variables prompted further investigation at a spatially resolved, subtype-specific level, as reported in the following section.

### Antipsychotic receptor occupancy dynamically modulates subtype-specific molecular-functional coupling in schizophrenia

To characterise how the neurotransmitter systems shape brain dynamics, we applied partial least squares (PLS) regression to map normative receptor density distributions [26], representing the cortical topography of dopaminergic, serotonergic, and other neuromodulatory receptors derived from Positron emission tomography (PET) and autoradiography in healthy populations, onto regional fluidity and ALFF. Analysis of PLS coefficients *β* revealed distinct patterns of molecular-functional coupling across clinical and neurobiological subtypes (Fig. 5a). A positive *β* coefficient indicates that cortical regions with higher local receptor density contribute more strongly to brain flexibility or local signal amplitude, that is, receptor-rich regions show greater fluidity or ALFF, whereas a negative *β* reflects an inverse relationship whereby regions densely expressing a given receptor are associated with reduced dynamic reconfiguration or signal power. Within the clinical data-driven clusters, the High Insight subtype exhibited significantly lower 5-HT_2*A*_ receptor-*β*_*fluidity*_ and lower D_1_ receptor-*β*_*ALFF*_ (*q* = 0.034) compared to the Low Insight group. The PANSS subtypes revealed even more pronounced differences: the HPHN subtype showed significantly elevated D_1_ receptor-*β*_*fluidity*_ (*q* = 0.008). The negative symptom distinction is represented by increased 5-HT_2*A*_ receptor–*β*_*ALFF*_ and 5-HT_6_ receptor– *β*_*ALFF*_ in LPHN versus LPLN, suggesting heterogeneous neuromodulatory architectures underlying functional dynamics across symptom profiles. In Fig. A8, the *β* coefficients within the entire dataset are presented.

**Fig. 5.**
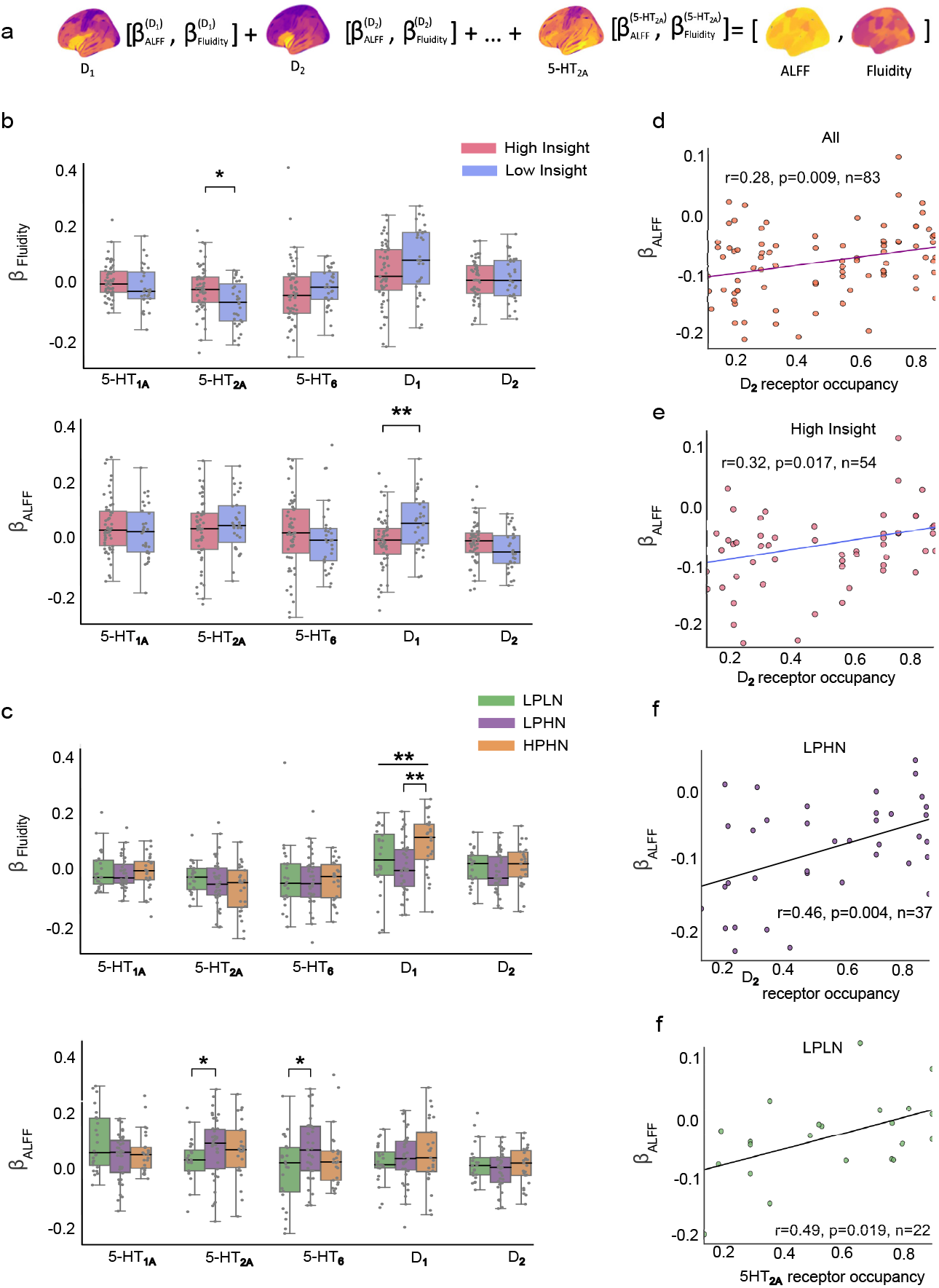
Molecular-functional coupling coefficients across clinical subtypes and their modulation by antipsychotic receptor occupancy. (a) PLS equation (b) *β* coefficients mapping normative receptor density distributions (5-HT_1*A*_, 5-HT_2*A*_, 5-HT_6_, D_1_, D_2_) onto regional fluidity (top) and ALFF (bottom) for High Insight versus Low Insight groups. High Insight showed significantly lower 5-HT_2*A*_–*β*_*fluidity*_ and lower D_1_–*β*_*ALFF*_ relative to Low Insight (^*^*q* < 0.05; ^**^*q* < 0.01). (c) PLS regression *β* coefficients PANSS-defined subgroups. HPHN exhibited elevated D_1_–*β*_*fluidity*_, and LPHN showed increased 5-HT_2*A*_– and 5-HT_6_–*β*_*ALFF*_ relative to LPLN, indicating heterogeneous neuromodulatory architectures across symptom profiles. Boxes represent interquartile range with median; whiskers extend to 1.5*×* IQR; individual data points overlaid. (d) Pearson correlations between estimated antipsychotic receptor occupancy and *β*_*ALFF*_ coefficients in the full cohort (*r* = 0.28, *p* = 0.009, *n* = 83) (e) Correlations in the High Insight subgroup (*r* = 0.32, *p* = 0.017, *n* = 54) (f) Correlatins in LPHN subgroup (*r* = 0.46, *p* = 0.004, *n* = 37), and the LPLN subgroup for 5-HT_2*A*_ occupancy (*r* = 0.49, *p* = 0.019, *n* = 22). Occupancy–*β* associations were predominantly observed for ALFF rather than fluidity, consistent with ALFF’s sensitivity to local, receptor-mediated neural fluctuations.

**Fig. 6.**
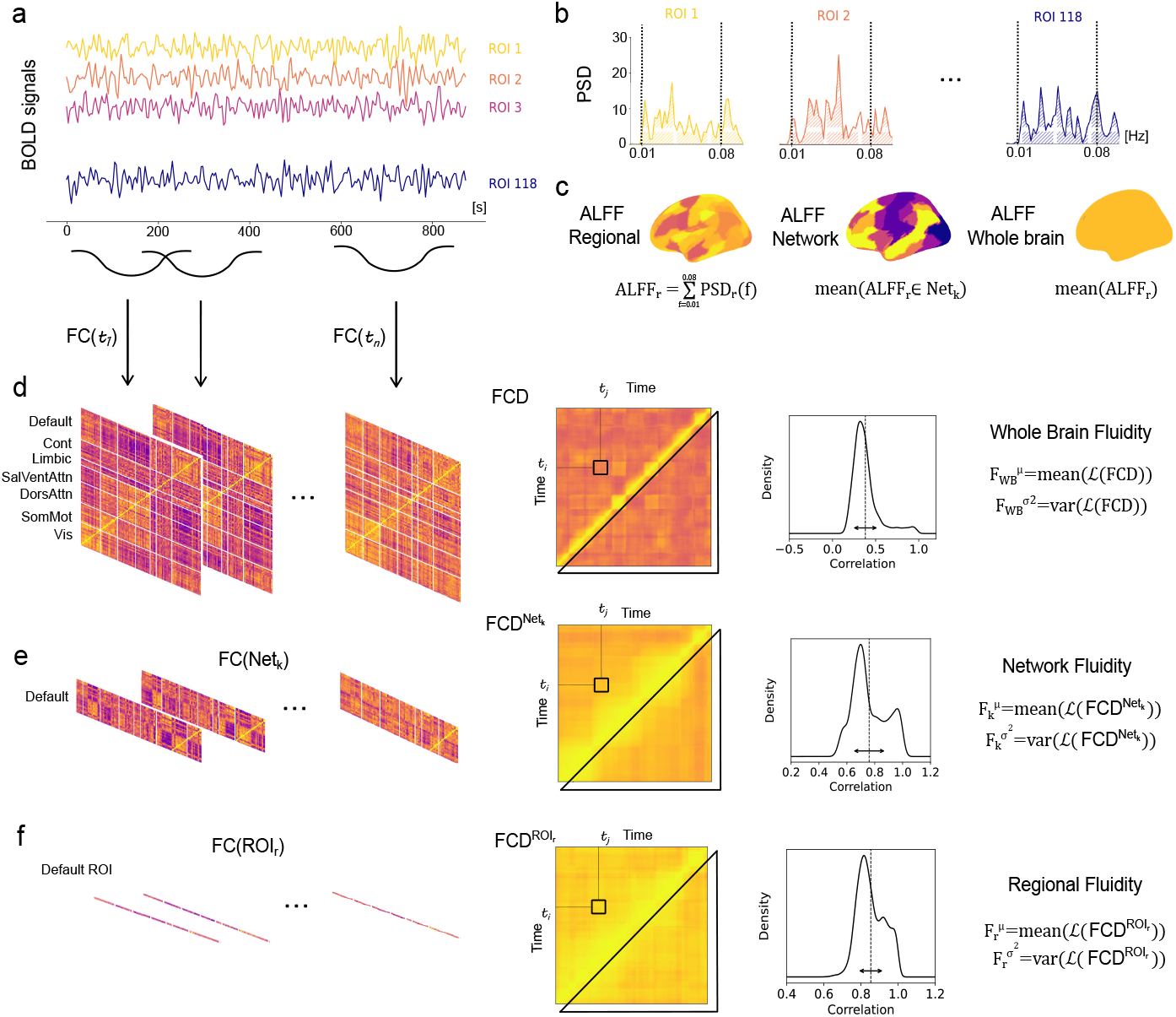
ALFF and Fluidity Feature extraction framework. (a) ROI BOLD time series extracted using the Schaefer 7-network, 100-parcel cortical atlas combined with 18 subcortical regions. (b) Power spectral density (PSD) estimated for each ROI using Welch’s method. (c) Regional ALFF computed as the shaded area within the predefined low-frequency band; network-level ALFF calculated as the sum of regional ALFF within each functional network; whole-brain ALFF defined as the mean of all regional ALFF values. (d) Whole-brain fluidity derived from sliding-window functional connectivity (FC): dynamic FC matrices were computed across time windows, FCD matrices were obtained by correlating FC matrices across windows, and fluidity was quantified as the mean and variance of the lower triangle of the FCD matrix. (e) Network-level fluidity calculated analogously, restricting FC and FCD computations to regions within a given functional network. (f) Regional fluidity computed using the same framework, restricted to a specific ROI.

Beyond subtype-level differences, we examined whether molecular-neural coupling (*β* coefficients) was dynamically modulated by antipsychotic exposure. Pearson correlations between estimated receptor occupancy and *β* coefficients revealed significant associations predominantly for *β*_*ALFF*_ rather than *β*_*fluidity*_ (Fig. 5 B). Across the full cohort, D_2_ receptor occupancy positively correlated with D_2_ receptor–*β*_*ALFF*_ (*r* = 0.27, *P* = 0.009, *n* = 83), with this relationship being the most pronounced in the LPHN subtype (*r* = 0.46, *P* = 0.004, *n* = 37). Within the High Insight group, D_2_ receptor occupancy similarly predicted D_2_ receptor–*β*_*ALFF*_ (*r* = 0.32, *P* = 0.017, *n* = 54). Additionally, 5-HT_2*A*_ receptor occupancy correlated with 5-HT_2*A*_ receptor–*β*_*ALFF*_ in the LPLN subtype (*r* = 0.49, *P* = 0.019, *n* = 22). All correlations are presented in Appendix A (Fig. A9, Fig. A11) for the full dataset and within subtypes, respectively. The predominance of *β*_*ALFF*_ over *β*_*fluidity*_ in these occupancy relationships likely reflects ALFF’s sensitivity to local, receptor-mediated fluctuations in neural activity, whereas fluidity captures global network reconfigurations that may be less directly constrained by regional receptor expression. Together, these findings indicate that clinical subtypes are characterised by distinct baseline patterns of molecular-functional coupling which are further dynamically modulated in a receptor- and region-specific manner by pharmacological exposure.

## Materials and Methods

### Data

The clinical dataset comprised 92 male patients with schizophrenia (aged 18–50 years), diagnosed according to DSM-IV criteria [27]. All participants were clinically stable for a duration of 6 months prior to the study. Demographic and medical variables were recorded for each participant, including age, weight, height, glycemia, smoking status, alcohol use, cannabis use, substance dependence, and illness duration. Symptom severity was assessed using the Positive and Negative Syndrome Scale (PANSS) [5], Calgary Depression Scale for Schizophrenia [28], Scale to Assess Unawareness of Mental Disorders (SUMD; all nine items included) [29], Global Assessment of Functioning (GAF) [30], and Clinical Global Impression (CGI) [31]. Antipsychotic medication type, dosage, and treatment-related side effects were available for 83 of 92 patients. Descriptive statistics are summarised in Table 1. Resting-state fMRI recordings of approximately 15 minutes duration were acquired for the patients; 5 were excluded due to insufficient data quality, yielding 87 usable scans.

### Clinical Data-Driven subtypes

By applying clustering algorithms to a 43-dimensional clinical feature space encompassing demographic, psychopathological, and physiological variables, including age, weight, height, blood glucose, illness duration, PANSS subscale scores (positive, PANP; negative, PANN; general psychopathology, PANG), all items from the SUMD and Calgary Depression Scale, CGI, GAF, and five antipsychotic-related side effects (concentration difficulties, asthenia, sedation, memory impairment, and depression). Prior to the analysis, missing values were imputed using k-nearest neighbours (*k* = 3) where each missing value was estimated as the mean of its three nearest neighbours [32], and all features were standardised (z-scored) to zero mean and unit variance. The optimal number of clusters was determined by evaluating internal validation metrics, including the Davies–Bouldin index [33], silhouette score [34], and elbow criterion. Across different clustering approaches, the most robust and parsimonious solution, consistent with Occam’s razor, was obtained using the k-means algorithm [35] *k* = 2, yielding two stable clusters termed High Insight and Low Insight. Principal Component Analysis [36] was performed for visualisation of the clinical data-driven subtypes, with the first two components explaining 28.5% of cumulative variance. The first principal component was primarily driven by SUMD item loadings, suggesting that illness insight was the dominant axis of inter-individual variation differentiating the two clusters.

### PANSS-defined subtypes

PANSS clusters were defined based on the positive and negative dimensions of the scale. Each score was computed as the sum of its respective subitems, *PANP* (positive) and *PANN* (negative). To capture clinically meaningful variation while maintaining balanced group sizes, we defined three groups: Low Positive–Low Negative (LPLN), Low Positive–High Negative (LPHN), and High Positive–High Negative (HPHN). A *Low Positive Score* was defined as a PANP sum ≤ 16, and a *Low Negative Score* as a PANN sum ≤ 22, based on score distribution within our dataset. According to these thresholds, only four participants fell into the High Positive–Low Negative (HPLN) group, which was underrepresented; therefore, two participants at the decision boundary were reassigned to the LPLN and two to HPHN groups to ensure balance.

### Pharmacology

Treatment data were available for 85 participants (81 with complete clinical data). Analyses were restricted to antipsychotic mediction. The majority of patients received antipsychotic monotherapy (82.2%), while a minority were treated with polytherapy (13.4%). Among monotherapy cases, aripiprazole, clozapine, olanzapine, risperidone (including long-acting injectable formulations), amisulpride, quetiapine, and haloperidol decanoate were prescribed. Zuclopenthixol and pipotiazine were excluded from pharmacological analyses owing to insufficient receptor-binding data. Receptor-binding affinities were defined from the literature data, focusing on dopaminergic, serotonergic, histaminergic, adrenergic, and muscarinic receptors [15, 37]. Receptor occupancy was estimated using a PK–PD framework linking administered dose to receptor affinity [38]. The pharmacokinetic component converted the daily dose into the estimated free plasma concentration available to cross the blood–brain barrier [39]:

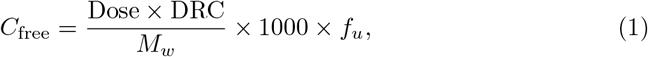

where *Dose* (mg/day), *DRC* (dose related concentration factor), *M*_*w*_ (molar mass), and *f*_*u*_ (unbound plasma fraction) are drug-specific parameters. Receptor occupancy for receptor *r* was computed using the Hill–Langmuir equation [38]:

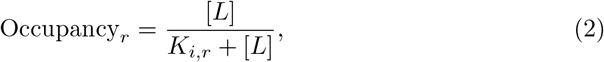

where [*L*] denotes the free drug concentration and *K*_*i,r*_ the affinity constant for receptor *r*. For polypharmacy, competitive binding was modelled using the Gaddum equation [40]:

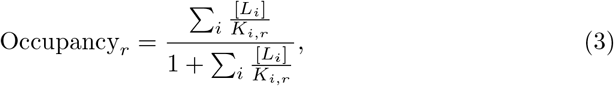

where [*L*_*i*_] and *K*_*i,r*_ represent the free concentration and affinity constant of the *i*-th drug at receptor *r*, respectively. Analyses focused primarily on dopaminergic (D_1−4_) and serotonergic (5-HT_1*A*_, 5-HT_2*A*_, 5-HT_2*C*_, 5-HT_6_) receptors given their central role in antipsychotic pharmacology. Additional receptor systems were included to account for adverse-effect profiles [41]. Although glutamatergic dysfunction is implicated in schizophrenia pathophysiology, it was not modelled directly due to limited pharmacological targeting by current antipsychotics [42].

### Neuroimaging Data

Functional images were acquired on a 3 T Siemens Verio scanner (CEMEREM, Marseille, France) using a 2D echo-planar imaging sequence (TR = 3600 ms, TE = 28 ms, flip angle = 90^°^, slice thickness = 2.5 mm, 50 axial slices, inter-slice spacing = 2.5 mm, in-plane resolution = 2.05 *×* 2.05 mm, matrix = 122 *×* 122, 100% phase FOV, partial Fourier = 0.75, parallel imaging factor = 2, bandwidth = 1413 Hz/pixel, phase encoding j–, total readout time = 47.8 ms) with a T:HEA;HEP coil. A reference volume was acquired for motion correction, and slices were acquired in ascending interleaved order. Images were converted to NIfTI format using *dcm2niix* (v1.0.20201224) [43]. After discarding patients who revoked consent and those whose scan duration was shorter than 15 minutes, data from 87 subjects were retained. Functional MRI data were preprocessed using fMRIPrep [44], a robust Nipype-based pipeline designed for BIDS-formatted datasets [45]. The anatomical reference used for preprocessing was each subject’s native T1-weighted (T1w) space, without transformation to a standard template. Functional images were corrected for head motion, susceptibility distortions, and slice timing, and were aligned to the corresponding T1w image using boundary-based registration [46].

Post-processing was performed in native T1w space using custom scripts based on NiLearn [47]. Confound regression was applied to each subject’s BOLD time series, removing six rigid-body motion parameters as well as mean signals from cerebrospinal fluid and white matter, as obtained from the fMRIPrep confounds file. No additional temporal derivatives, quadratic terms, or global signal regression were applied. Regression was performed using ordinary least squares as implemented in Nilearn’s cleaning function. The signals were further band-pass filtered using a 6th-order Butterworth IIR filter (0.01–0.1 Hz), following standard denoising practices [48]. This preprocessing strategy preserved the original spatial and temporal resolution while controlling for primary sources of non-neuronal variance. Time series were extracted from denoised BOLD data using the Schaefer 7-network parcellation with 100 cortical regions [49], supplemented with 18 subcortical regions from a corresponding subcortical atlas [50]. The resulting 118 regional time series were used for downstream analyses.

### Neuroimaging Feature Extraction

#### Static and Dynamic Functional Connectivity

Static functional connectivity (FC) was estimated using Pearson correlation between regional time series via the nilearn ConnectivityMeasure implementation [47]. Network-level segregation and integration were computed by averaging within-network and between-network connectivity strengths, respectively [51].

Functional connectivity dynamics were derived using a sliding-window approach (window length *L* = 15 TRs, step size *S* = 2 TRs). Windowed correlation matrices were used to compute the mean and variance of time-resolved segregation and integration. FCD matrices were obtained by correlating vectorised lower-triangular connectivity patterns across windows [52]. Intra- and inter-network fluidity were quantified as the variance and mean of nonoverlapping upper-triangular FCD values, reflecting the stability and flexibility of connectivity configurations over time [53, 54]. Regional fluidity was calculated using the same approach as depicted in Fig. 6.

#### Spectral Features

Spectral properties were estimated using Welch’s power spectral density method. The amplitude of low-frequency fluctuations (ALFF) was computed as the square root of the integrated power within 0.01–0.08 Hz [55]. Fractional ALFF (fALFF) was calculated as the ratio of low-frequency power to total spectral power [56]. Additional band-limited power measures were derived by integrating spectral density within predefined frequency ranges.

#### Phase Synchronisation and Metastability

The instantaneous phase of each regional BOLD time series was extracted using the Hilbert transform. Global phase synchronisation was quantified using the Kuramoto order parameter (KOP), computed from the complex phase representation across all regions at each time point [57, 58]. The temporal mean of the KOP reflects the overall level of whole-brain synchrony, whereas its temporal variance was used as an index of metastability, capturing the degree to which the system dynamically transitions between more and less synchronised states over time [59, 60].

### Machine Learning Analysis

To differentiate between the clinical data-driven and PANSS-defined subtypes, we conducted a machine learning analysis and explored the most important features that aid the subtype distinction. We applied supervised learning algorithms to the high-dimensional dataset of functional neuroimaging features. For 86 patients, we included all the regional, network, and whole-brain features and applied a classification pipeline for a binary classification between High Insight and Low Insight patients, and multiclass classification for the PANSS clusters, LPLN, LPHN, and HPHN. All features were first standardised using `StandardScaler` to ensure comparable scales across predictors [61].

To reduce dimensionality and retain the most informative features, a univariate feature selection step (`SelectKBest` with ANOVA *F* -score) was applied [62], with the number of selected features optimised for the SVM classifier. Hyperparameter tuning was performed using grid search within a nested cross-validation (nested-CV) framework [63]. An outer 3-fold stratified cross-validation provided unbiased estimates of predictive performance, while an inner 5-fold stratified CV optimised hyperparameters. Model performance was quantified using accuracy, balanced accuracy, F1-score (macro for multiclass), and ROC-AUC [64], allowing robust comparison across classifiers and target variables. Feature importance was assessed via permutation importance [65], which measures the decrease in model performance upon random shuffling of each feature, and SHAP (SHapley Additive exPlanations) values [66], providing model-agnostic, per-feature contributions to individual predictions. Both approaches allow identification of key features driving classification outcomes, supporting biological interpretability.

#### Classification Pipeline and Hyperparameter Optimisation

For the classification framework, support vector machine (SVM)[67] was implemented within a supervised learning pipeline. Each pipeline incorporated a *z*-score normalisation step using `StandardScaler` [61] to account for differences in feature scales, followed by univariate feature selection via `SelectKBest` [62] to reduce dimensionality and retain the most informative predictors. For the binary classification task, the optimal configuration consisted of retaining 816 features and training a linear SVM with a regularisation parameter *C* = 0.01, employing class weighting to mitigate class imbalance, and enabling probability estimation for subsequent analyses. The best pipeline for classifying the fluidity features was achieved by selecting 60 of them using a linear SVM with *C* = 1. In contrast, for the multiclass classification task, the best-performing model was achieved with a substantially more constrained feature set of 20 predictors, coupled with a linear SVM with stronger regularisation (*C* = 10), again with balanced class weighting and probabilistic outputs. These results highlight classification-specific differences in the optimal feature space dimensionality and regularisation strength, suggesting that while binary discrimination benefits from a broader representation of features, multiclass differentiation relies on a more selective and tightly regularised subset of features to achieve robust performance [67, 68].

#### Feature Importance and Distribution Analysis

To characterise which neuroimaging feature types, brain networks, and spatial scopes (whole-brain, hemispheric, network-level, regional) contributed most to classification, we employed a multi-layered importance analysis fully embedded within the nested-CV framework. Comparison of four complementary distributions, namely the null distribution, the ANOVA-selected distribution, permutation importance, and SHAP importance, allowed identification of categories that were overrepresented relative to the chance, indicating a genuine predictive signal rather than bias from the candidate feature set. As a baseline reference, we computed the proportional representation of each feature category (feature type, brain network, spatial scope) across the *entire* feature matrix prior to any selection or modelling. This null distribution reflects the expected category proportions under random sampling from the full pool, and serves as the reference against which all downstream distributions are compared. Within the inner loop of each outer fold, the ANOVA *F* -test (`SelectKBest`) retained the *k*^*^ features most univariately associated with the class label [62]. To obtain a consensus set of robustly relevant features, we computed the *strict intersection* across all outer-fold best estimators: only features selected in *every* fold were retained. To investigate the contribution of individual features to the model performance, we quantified feature importance using permutation importance [65], with balanced accuracy as the scoring function and 20 permutation repetitions per feature per fold. Because the present dataset is small, setting aside a dedicated held-out test set would have substantially reduced both training and evaluation sample sizes, potentially destabilising importance estimates. We therefore adopted a *train-set permutation importance* strategy, which estimates the degree to which each feature is relied upon by the fitted model referred to here as **model reliance** rather than its out-of-sample generalisation contribution [69]. Fold-level importance scores were averaged across the three outer-fold estimators to yield a stable, cross-validated estimate of model reliance. Permutation importance highlights features that are indispensable for overall predictive accuracy; it may, however, underestimate the importance of redundant features when overlapping predictors can compensate for one another. Absolute importance values were aggregated by feature category, brain network, and spatial scope, and then normalised to unit-sum proportions. This produced a distribution that is directly comparable to the null and ANOVA distributions. SHAP (SHapley Additive exPlanations) values [66], which provide an analytically exact decomposition for linear models [70], were obtained by passing the full dataset through the preprocessed feature space of each fold’s estimator. Mean absolute SHAP values were computed per feature, averaged across folds, aggregated by category, and normalised to proportions. The combination of permutation importance and SHAP values offers complementary insights: permutation importance highlights features indispensable for overall predictive accuracy, while SHAP emphasises the consistency and directionality of feature effects across samples, providing theoretically grounded attribution at the feature level [66, 69]. Concordance across all three model-derived distributions strengthens the interpretation that identified features reflect robust, method-invariant neural signatures of the classification target [65, 66].

### Structural Equation Modeling

To examine the interrelationships between pharmacological markers, brain connectivity, and clinical outcomes, we specified a structural equation model comprising three latent factors [71, 72]. The neuroimaging factor (*Latent Neuro*) was defined using the most statistically significant rs-fMRI features identified after false discovery rate correction [73]. The pharmacological factor (*Latent Pharmo*) was indexed by receptor-related measures (D2, D3, and 5-HT_2*A*_), reflecting dopaminergic and serotonergic receptor profiles. The clinical domain (*Latent Clinic*) was characterised by symptom severity measures (SUMD, PANP, and PANN), capturing complementary dimensions of psychopathology [74]. Structural paths were specified to test both direct and indirect associations between pharmacological, neurobiological, and clinical domains [72]. Model parameters were estimated using maximum likelihood with latent variables scaled to unit variance, and mean structures were included [71, 75]. Statistical inference was performed using nonparametric bootstrap resampling (2,000 iterations) to obtain robust standard errors and bias-corrected confidence intervals for all parameter estimates [76]. Model adequacy was evaluated using standard goodness-of-fit indices [77], and standardized estimates are reported throughout.

### Partial Least Squares and Pharmacological Relations

To examine the multivariate relationship between regional neurotransmitter receptor distributions and functional brain dynamics, we applied subject-level partial least squares regression [78]. PLS was chosen because it allows simultaneous modelling of multiple correlated outcome variables, capturing latent components that represent shared covariance between predictors and outcomes, which is particularly suitable for jointly analysing fluidity and ALFF. Receptor density maps were parcellated using the 100-region Schaefer atlas [49] and z-scored across parcels within each subject to obtain predictor matrices (**X**), standardising values to ensure comparability across regions and subjects. Regional fluidity and ALFF were jointly modelled as multivariate outcomes (**Y**), allowing the PLS model to capture patterns in functional dynamics that are shared across these complementary measures. For each subject, a two-component PLS model was estimated using singular value decomposition, yielding receptor-specific *β* coefficients reflecting the strength and direction of association between receptor distributions and functional brain dynamics, as well as predicted functional values for each parcel. Model performance was quantified by computing the Pearson correlation between observed and predicted functional measures across parcels, providing an intuitive measure of how well receptor distributions explain individual functional profiles. Parcel-wise measures are inherently spatially autocorrelated, meaning that nearby brain regions tend to have similar functional values [79, 80], and our current approach does not explicitly account for this dependency, which may inflate apparent statistical associations or affect interpretability [81]. Nevertheless, summarising *β* coefficients across subjects using medians and interquartile ranges provides a robust description of central tendency and inter-subject variability without assuming parametric distributions. To explore differences between clinically derived subtypes, pairwise differences in PLS *β* coefficients were assessed using two-sided Mann–Whitney U tests for clinical data-driven clusters; for PANSS-defined groups, overall group effects were evaluated using Kruskal–Wallis tests, followed by post hoc pairwise Mann–Whitney U tests with false discovery rate (FDR) correction. These non-parametric tests were selected because *β* coefficients may not satisfy the assumptions of parametric tests, and to provide interpretable, distribution-free effect estimates across heterogeneous clinical subgroups. Finally, to investigate pharmacological modulation of brain–molecular coupling, subject-level PLS *β* coefficients were correlated across subjects with corresponding receptor occupancy values derived from pharmacological data, assessing how individual variation in receptor engagement by antipsychotic medication influences the association between receptor distributions and functional brain dynamics [26], and providing insight into potential neuromodulatory mechanisms underlying functional alterations in schizophrenia [82].

## Discussion

We developed an integrative multimodal framework that combines data-driven clinical subtyping with resting-state fMRI and PK–PD estimated occupancy to characterise the neurobiological and pharmacological heterogeneity of schizophrenia. Our results establish a reproducible link between illness insight, dynamic network flexibility, and receptor-level pharmacology. Moreover, we identified regional fluidity as a candidate neural biomarker capable of stratifying patients into clinically and biologically distinct subtypes.

### Illness insight as the principal axis of clinical variation

Unsupervised clustering of a 43-dimensional clinical feature space yielded two robust subtypes whose separation was principally organised along an axis of illness insight. Crucially, positive and negative symptom burden co-varied with this axis: patients with attenuated syndromal severity tended to preserve greater metacognitive awareness, whereas those carrying higher symptom loads exhibited markedly impaired illness insight. This structure was not imposed by the algorithm but emerged from data geometry, lending the identified subtypes validity as clinically meaningful entities rather than statistical artefacts [83, 84]. The primacy of insight in organising inter-patient variability is consistent with models that implicate frontoparietal and default mode network dysfunction in metacognitive monitoring [85]. Insight has long been recognised as a partially independent dimension of psychopathology [86]; yet here it emerged as the dominant axis of clinical variation, suggesting it may be more tractable as a stratification variable than the positive–negative dichotomy alone. This aligns with precision psychiatry frameworks that advocate moving beyond DSM categorical boundaries toward richer, biologically grounded representations of psychopathological state-space [24, 87].

### Regional fluidity discriminates insight-based subtypes

Across both SUMD- and PANSS-defined groups, rs-fMRI features provided a discriminative signal that exceeded the null distribution defined by feature category composition, confirming the relevance of macroscale brain dynamics to schizophrenia phenomenology [17, 18]. Critically, the most informative features differed between subtyping approaches. Regional *fluidity* —a measure of temporal variability in functional connectivity dynamics derived from the variance of FCD matrices [52, 53]—was the principal discriminative measure for insight-based subtypes, with 48 of 100 cortical regions showing significantly higher fluidity in High Insight patients after FDR correction. The largest effect sizes was observed in the Default Mode Network (mean |*d*| = 0.54) and Somatomotor Network (mean |*d*| = 0.61), regions associated with self-referential processing and motor predictive coding, respectively [88, 89]. This finding extends prior reports of reduced dynamic functional connectivity in schizophrenia [90, 91] and is consistent with evidence linking impaired insight specifically to the altered resting-state functional connectivity patterns [92], connecting metacognitive deficits to restricted network flexibility. Theoretically, fluidity can be understood as an index of the brain’s capacity to explore its functional state-space [93, 94], a capacity that may be necessary for accurate belief updating about one’s own mental states. The dissociation we observe with fluidity being most discriminative for insight subtypes while ALFF was more informative for PANSS subtypes, suggests that distinct aspects of psychopathology are preferentially encoded in different dimensions of spontaneous brain activity. Moment-to-moment fluctuations in network configuration may be particularly relevant to metacognitive function, whereas local neural excitability patterns, as captured by ALFF [55], may be more tightly coupled to syndromal symptom severity and dopaminergic dysregulation [95, 96]. All rs-fMRI data were acquired under ongoing antipsychotic treatment, and the observed functional differences are expected to reflect a combination of disease-related pathological alterations and treatment-induced neuromodulatory effects. Despite receptor occupancy representing a global whole-brain estimate, its correlation with regional fluidity allowed us to identify spatially heterogeneous patterns of pharmacological modulation across the cortex. When interpreting these results, it is important to have in mind the modulation constrained by underlying receptor density architectures, which is not considered when computing receptor occupancy.

Support vector machine classification using the full 819-dimensional neuroimaging feature space (accuracy 0.76 ± 0.07) and a compact fluidity-dominating subset (SVM 2: accuracy 0.67 ± 0.12) confirmed that regional fluidity alone retains substantial discriminatory information. The lower but above-chance performance of SVM 3 for PANSS-defined groups (accuracy 0.58 ± 0.09) reflects the greater neurobiological overlap across PANSS-defined groups, consistent with evidence that symptom severity dimensions do not map directly onto distinct neural substrates [9].

#### Pharmacological effects are subtype-dependent

Receptor occupancy–symptom associations were sparse and weak in the full cohort but emerged as substantially stronger and more widespread within specific subtypes. The High Positive–High Negative group exhibited the highest density of significant correlations (|*r* |≈ 0.4–0.6), encompassing multiple dopaminergic and serotonergic receptor–symptom pairs. The Low Positive–Low Negative subgroup, by contrast, showed only limited pharmacological–clinical coupling, suggesting a relatively attenuated receptor–symptom interface in this low-burden group. These observations directly parallel the long-recognised inter-individual variability in antipsychotic response [3, 97], which is mainly attributed to pharmacokinetic differences. Inter-individual variability in antipsychotic response has traditionally been attributed to pharmacokinetic differences, such as how drugs are metabolised and distributed across patients. Our results suggest that this view may be incomplete: the relationship between receptor occupancy and clinical outcome appears to differ across neurobiological subtypes. In other words, treatment variability may reflect not only pharmacokinetics but also pharmacodynamic heterogeneity, with the brain’s response to receptor engagement varying between subtypes. This reframing resonates with precision medicine frameworks that seek to align pharmacological targets with patient-specific neurobiological profiles [4, 24, 98]. The selective association of 5-HT_2*A*_ occupancy with positive symptom measures in the LPHN subtype, together with the broader D_2_ occupancy associations in HPHN patients, further suggest subtype-specific patterns of receptor–symptom coupling that are not apparent at the population level [99].

#### Neuroimaging mediates the pharmacology–symptom relationship

Structural equation modelling formalised the hypothesis that neuroimaging occupies a mediating role on the pathway from receptor occupancy to clinical outcomes. The fitted model demonstrated excellent global fit [100, 101], and the latent Neuroimaging factor was a significant predictor of latent Clinical severity (*β* = −0.26, *p* = 0.041), indicating that greater or more intact neurofunctional dynamics corresponded to lower symptom burden. Notably, the direct path from the latent Pharmacology factor to Neuroimaging did not reach significance, which we attribute primarily to a spatial resolution mismatch: receptor occupancy was operationalised as a whole-brain scalar quantity [102], whereas neuroimaging features were derived at the regional level [49]. This scale discrepancy is a fundamental limitation of pharmacokinetic– pharmacodynamic approaches that collapse distributed receptor engagement into a single global estimate. Positron emission tomography measures of regional receptor occupancy [103, 104] or pharmacokinetic modelling with spatially resolved compartmental parameters would offer the resolution required to detect pharmacological effects on regional dynamics. The present SEM findings should therefore be understood as providing a lower-bound estimate of the neuroimaging-mediated pharmacological influence, rather than ruling out such an effect.

#### Molecular architecture shapes subtype-specific functional dynamics

Partial least squares regression mapping normative neurotransmitter receptor density distributions [26] onto regional fluidity and ALFF revealed that these two functional metrics are coupled to distinct molecular architectures. In the clinical data-driven subtypes, High Insight patients exhibited significantly lower 5-HT_2*A*_ receptor–*β*_*fluidity*_ and lower D_1_ receptor–*β*_*ALFF*_ relative to Low Insight patients (*q* < 0.05). Among PANSS-defined groups, the HPHN subtype showed elevated D_1_ receptor–*β*_*fluidity*_, whereas the LPHN subtype displayed increased 5-HT_2*A*_ and 5-HT_6_ receptor–*β*_*ALFF*_ relative to LPLN—indicating heterogeneous neuromodulatory architectures underlying symptom profiles [14, 15]. Beyond group-level baseline differences, antipsychotic receptor occupancy dynamically modulated these molecular–functional associations. D_2_ occupancy positively predicted *β*_*ALFF*_ in the full cohort (*r* = 0.27, *p* = 0.009) and more strongly within the LPHN (*r* = 0.46, *p* = 0.004) and High Insight (*r* = 0.32, *p* = 0.017) subgroups. The predominance of *β*_*ALFF*_ over *β*_*fluidity*_ in these occupancy relationships is mechanistically interpretable: ALFF indexes local neural excitability that is directly sensitive to receptor-mediated synaptic modulation, while fluidity reflects distributed network reconfiguration that is only indirectly constrained by regional receptor density as suggested by prior work on neuromodulatory control and whole-brain dynamics [93, 105]. This dissociation has translational implications—ALFF may constitute a more sensitive pharmacodynamic readout for tracking treatment response in clinical trials [18, 96].

### Limitations

Several constraints limit the interpretation of these findings. The cohort comprised 92 male patients, restricting both statistical power and generalisability given established sex differences in schizophrenia symptom expression, illness course, and pharmacological response [106, 107]. Although adequate for exploratory analyses using interpretable, low-complexity models, this sample size precludes more flexible deep learning or high-dimensional latent variable approaches; replication in larger, sex-balanced, independent cohorts is essential [108].

Preprocessing choices, particularly the decision against global signal regression, substantially influenced the spatial and temporal structure of extracted features and their interpretation, a well-recognised challenge in the field [48, 109]. Systematic evaluation across preprocessing pipelines and adoption of community harmonisation standards are needed to improve reproducibility [110]. From a pharmacological standpoint, our analyses focused on antipsychotic medications. However, some patients were concurrently prescribed antidepressants, which can act on overlapping receptor systems and whose contributions were not explicitly modelled but are sensitive to pharmacogenetic variability, particularly cytochrome P450 polymorphismsm which was not accounted for owing to the absence of genotyping data [39, 111]. Integrating CYP1A2 phenotyping, a key determinant of clozapine and olanzapine exposure would reduce residual variance in occupancy estimates and improve their biological fidelity. The SEM represents a deliberate simplification and should be understood as capturing a subset of what are likely far more intricate, mutually reinforcing relations among latent clinical, neuroimaging, and pharmacological constructs [101, 112]. Furthermore, the use of normative receptor maps derived from healthy populations as a proxy for individual molecular architecture is an important caveat. Disease-related alterations in receptor expression, including D_2_ upregulation and 5-HT_2*A*_ changes documented in schizophrenia post-mortem and PET studies [12, 82] may render normative maps imperfect surrogates. Patient-specific PET receptor quantification would substantially improve the resolution of molecular-functional coupling analyses in future work [103, 104]. Finally, parcel-wise measures are inherently spatially auto-correlated [79, 80], and our PLS analyses do not explicitly correct for this dependency, which may inflate apparent statistical associations.

### Future directions

These limitations collectively underscore the need for large-scale, purpose-built multimodal datasets integrating longitudinal clinical assessments, structural and functional MRI, PET-based receptor quantification, and pharmacogenomic profiling, ideally across multiple sites with harmonised acquisition protocols [110]. Longitudinal designs would further clarify the directionality of the relationships described here, distinguishing state-dependent changes in functional dynamics from trait-level neurobiological vulnerabilities that antedate illness exacerbation. Incorporating computational models of neuromodulation, such as dynamic mean-field or neural mass models constrained by receptor density maps [93, 113] would additionally allow formal testing of mechanistic hypotheses about how antipsychotic receptor engagement reshapes macroscale network dynamics across subtypes.

## Conclusion

Schizophrenia’s clinical and neurobiological heterogeneity has long obstructed the identification of reliable biomarkers and personalised treatment strategies. By integrating clinical, neuroimaging, and pharmacological data within a unified framework, the present work demonstrates that stratifying patients on the basis of illness insight, a clinically underutilised dimension, yields neurobiologically and pharmacologically coherent subtypes. Among the neuroimaging features examined, the fluidity of regional functional connectivity emerged as the strongest discriminator of insight-based subtypes, capturing moment-to-moment reconfigurations of network topology that were consistently attenuated across all functional networks in Low Insight patients. In contrast, PANSS-based groupings did not converge on a single defining neuroimaging signature, reinforcing the value of insight as an organising principle for subtype discovery and underscoring that syndromal severity and neural dynamics need not co-vary isomorphically.

Multivariate classification confirmed the discriminative power of rs-fMRI, achieving robust performance even when restricted to the most informative fluidity features, supporting their candidacy as functional biomarkers. Partial least squares analyses revealed that the coupling between normative receptor density architectures and regional brain dynamics is systematically distinct between subtypes, with serotonergic and dopaminergic systems exhibiting different associations with fluidity and ALFF depending on clinical profile. D_2_ and 5-HT_2*A*_ receptor occupancy selectively modulated ALFF-based molecular-functional coupling, implicating these systems as key pharmacodynamic levers whose effects are contingent on the patient’s underlying neurobiological subtype. Structural equation modelling provided formal evidence that neural dynamics mediate the relationship between receptor-level pharmacology and clinical symptom expression, positioning neuroimaging as a mechanistic bridge between molecular and phenomenological levels of analysis [16, 93, 105].

Together, these findings indicate that integrating clinical stratification, dynamic functional neuroimaging, and pharmacological characterisation can uncover reproducible, biologically meaningful schizophrenia subtypes, identify candidate functional biomarkers, and illuminate the receptor-level mechanisms through which antipsychotic treatment exerts its differential clinical effects. This multimodal strategy offers a concrete roadmap toward precision psychiatry in schizophrenia: one in which pharmacological targets are matched to patient-specific neurobiological profiles, and neuroimaging signatures serve as quantitative readouts of therapeutic engagement [4, 24, 98].

## Data Availability

All data produced in the present study are available upon reasonable request to the authors

## Acknowledgements

This work was supported by the A*MIDEX foundation (Aix-Marseille University Initiative of Excellence) through the 2021 Interdisciplinarity Call for Projects, under the project “REPORTS” (bRain modEling and PharmacOclinical Response To Schizophrenia).

This project was carried out within Aix-Marseille University and its affiliated institutions, including the Institut de Neurosciences des Systèmes (Inserm UMR 1106), the Centre for Magnetic Resonance in Biology and Medicine (CRMBM UMR 7339), and the Academic Department of Psychiatry at AP-HM (Schizophrenia Expert Center, FondaMental Foundation).

In addition this research has received funding from the European Union’s Horizon Europe Programme under the Specific Grant Agreement No. 101147319 (EBRAINS 2.0 Project) and No. 101137289 (Virtual Brain Twin Project). The funders had no role in study design, data collection and analysis, decision to publish, or preparation of the manuscript.

All authors report no biomedical financial interests or potential conflicts of interest.

## Appendix A Supplementary

**Fig. A1.**
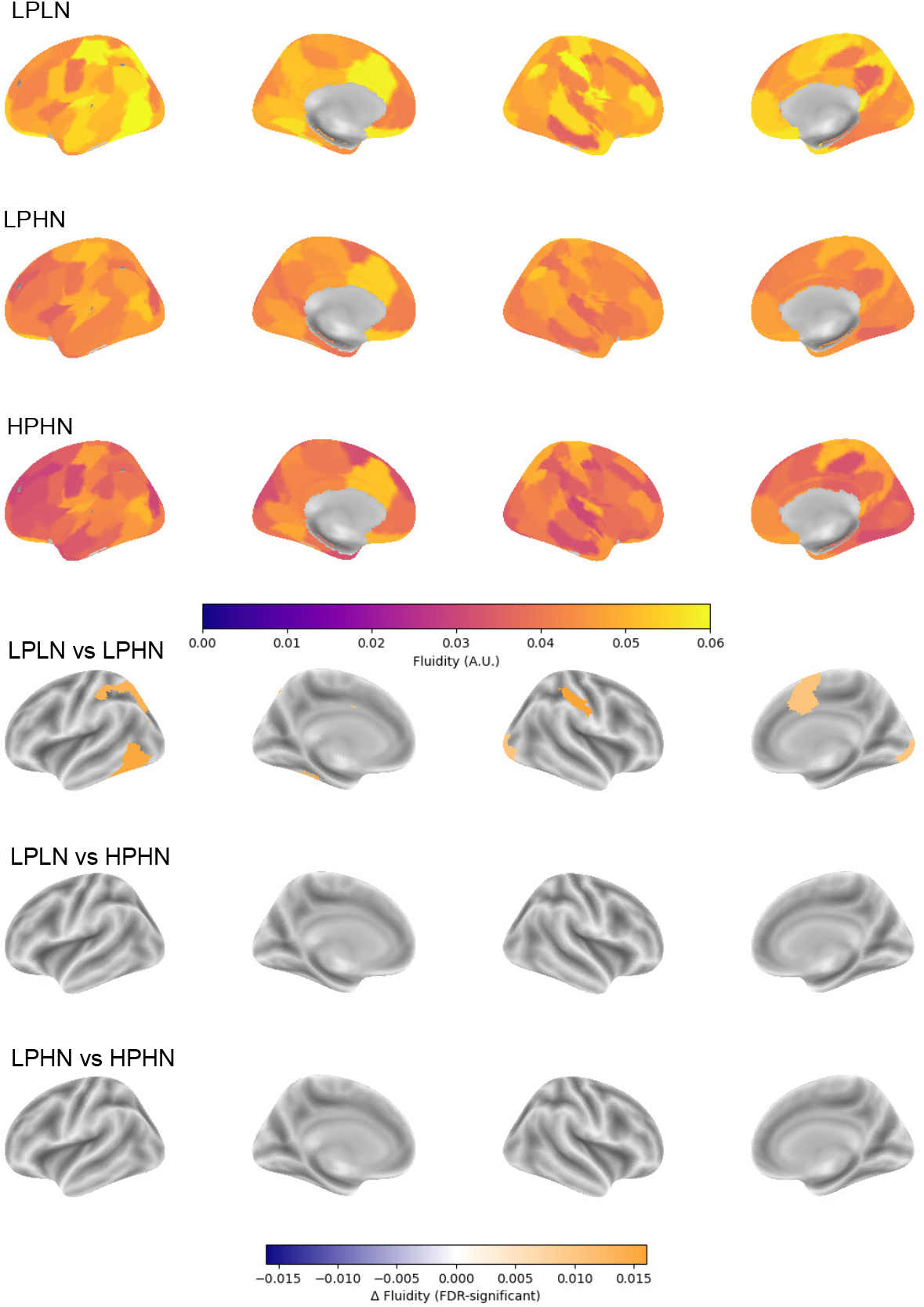
Mean regional fluidity in PANSS-defined clusters. Gradient conserved with fewer symptoms exhibited higher regional fluidity, whereas the highest symptom profiles exhibited lower fluidity. Kruskal-Wallis test did not show significance between groups (*p* < 0.05), post-hoc Mann–Whitney U test, FDR corrected, showed significant regional fluidity between LPLN and LPHN, portraying the difference in manifestation of negative symptoms in the brain dynamics.

**Fig. A2.**
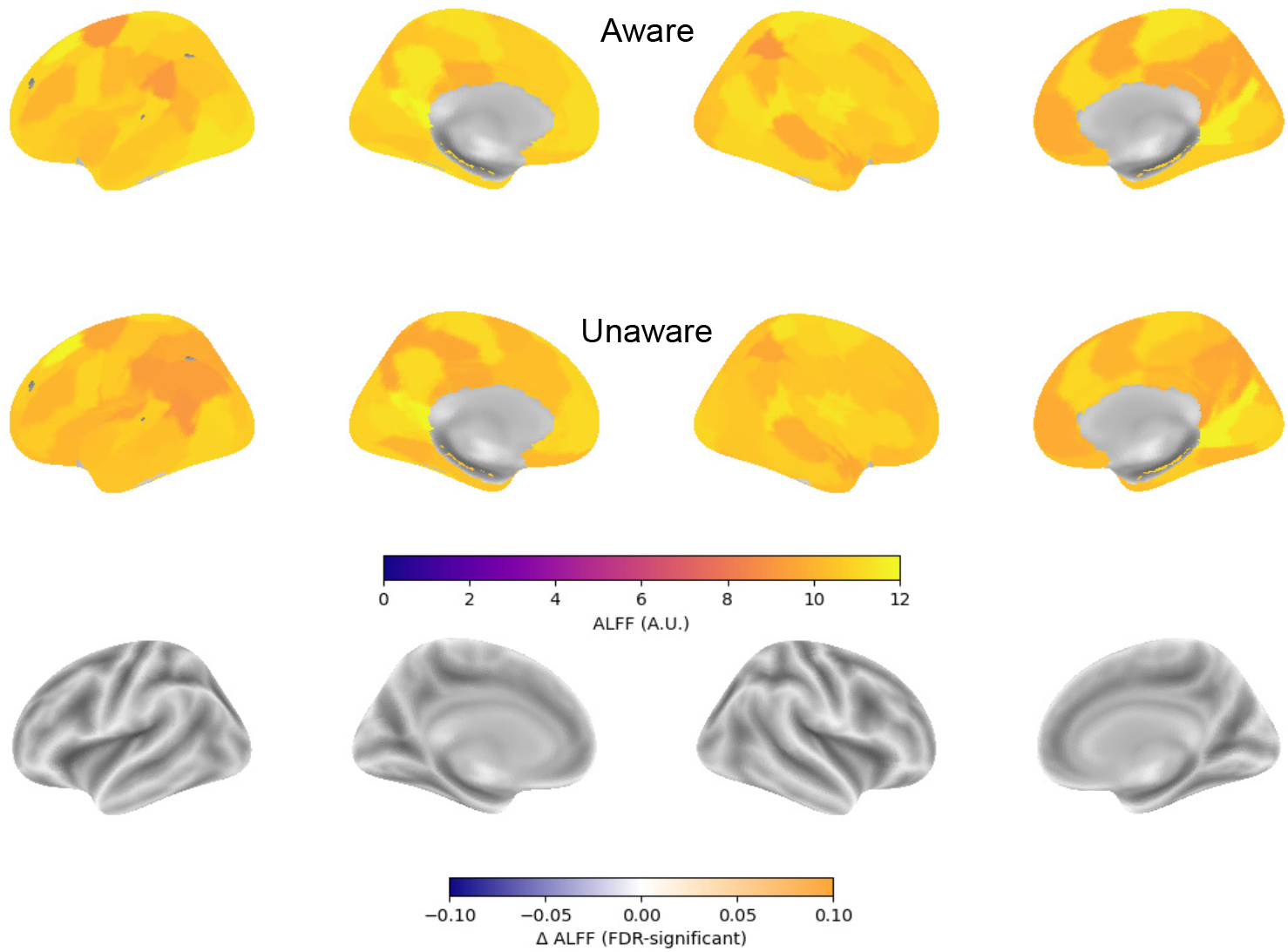
Mean regional ALFF in Aware or High Insight vs Unaware or Low Insight patients. No significant intergroup differences, Mann–Whitney U test (*p* < 0.05)

**Fig. A3.**
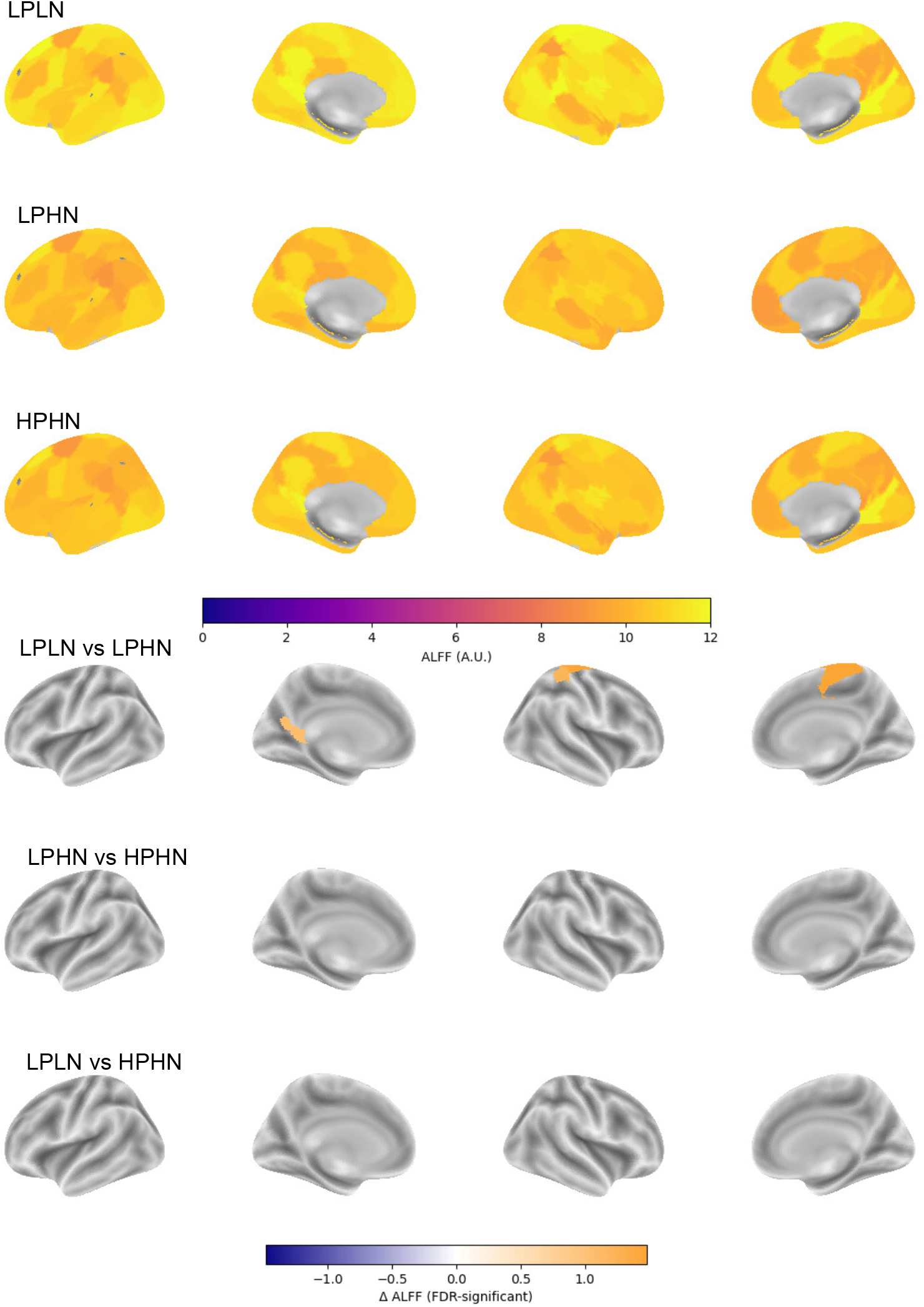
Mean regional ALFF in PANSS-clusters. Kruskal-Wallis test did not show significance between groups (*p* < 0.05), post-hoc Mann–Whitney U test FDR corrected, showed significant regional fluidity between LPLN and LPHN, portraying the difference in manifestation of negative symptoms in ALFF as well.

**Fig. A4.**
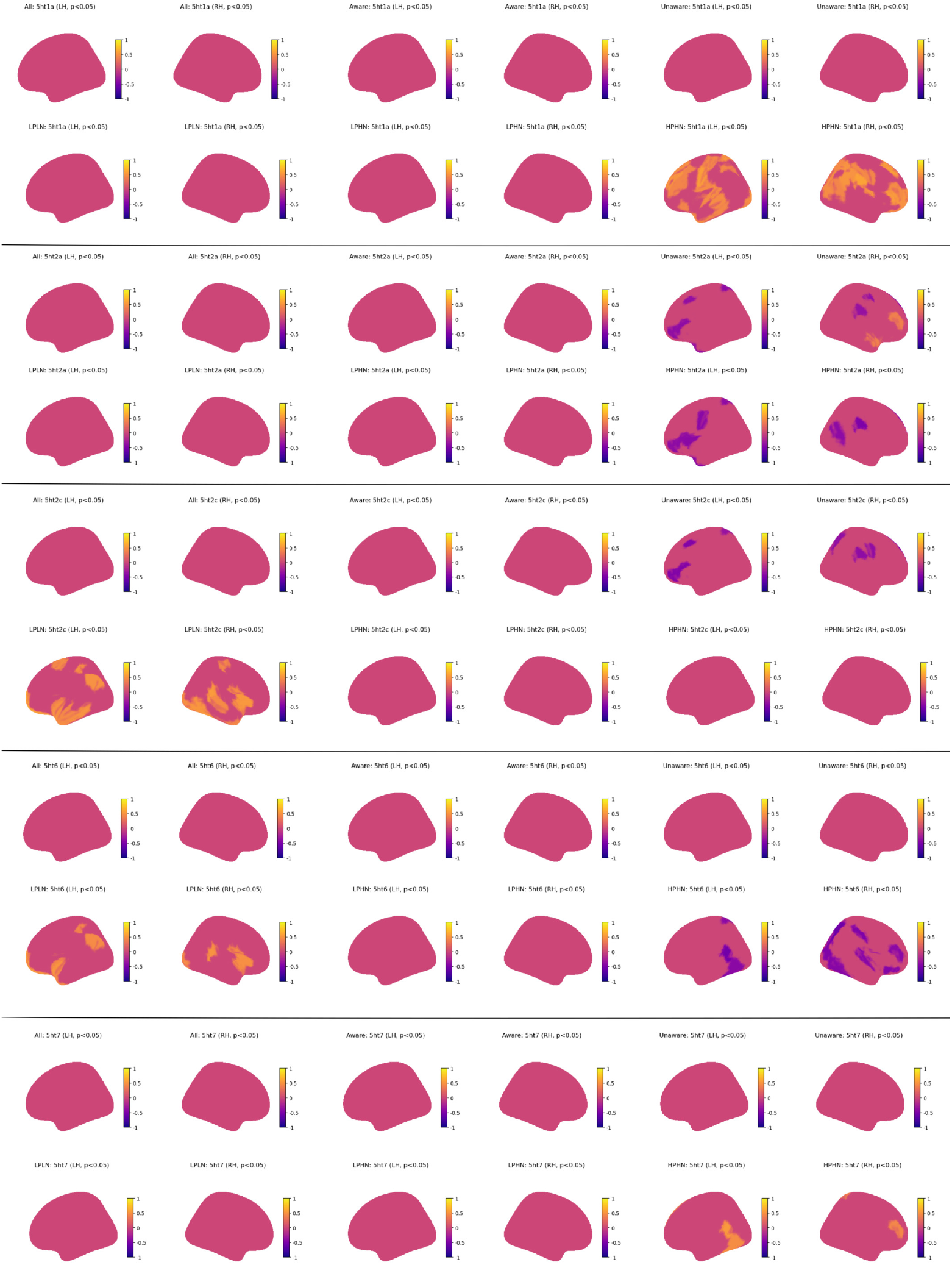
Significant correlations of regional fluidity and receptor occupancy. Orange regions indicate positive correlations and purple indicating negative correlations of the serotonergic receptors.

**Fig. A5.**
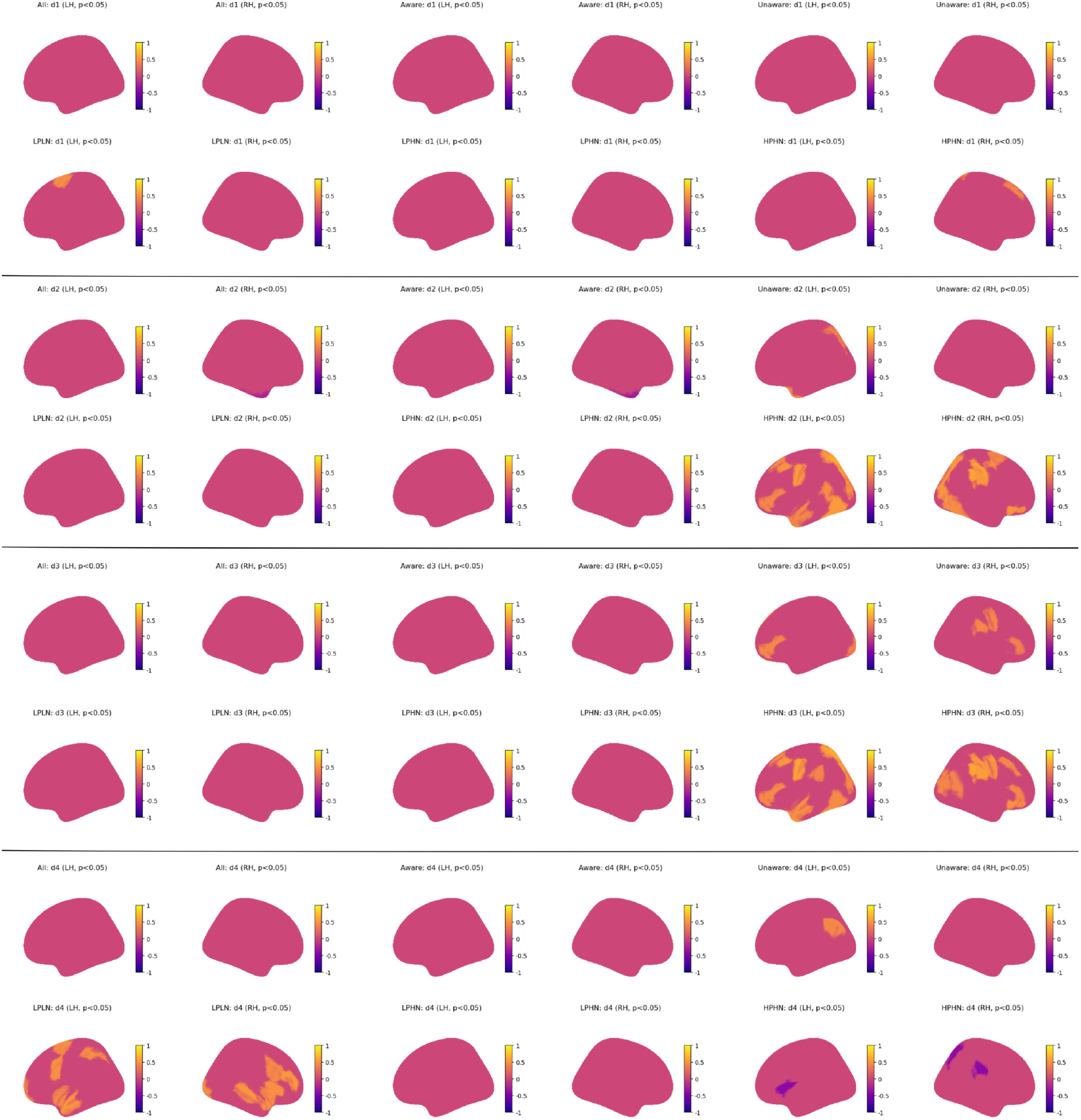
Significant correlations of regional fluidity and receptor occupancy. Orange regions indicate positive correlations and purple indicating negative correlations of the dopaminergic receptors.

**Fig. A6.**
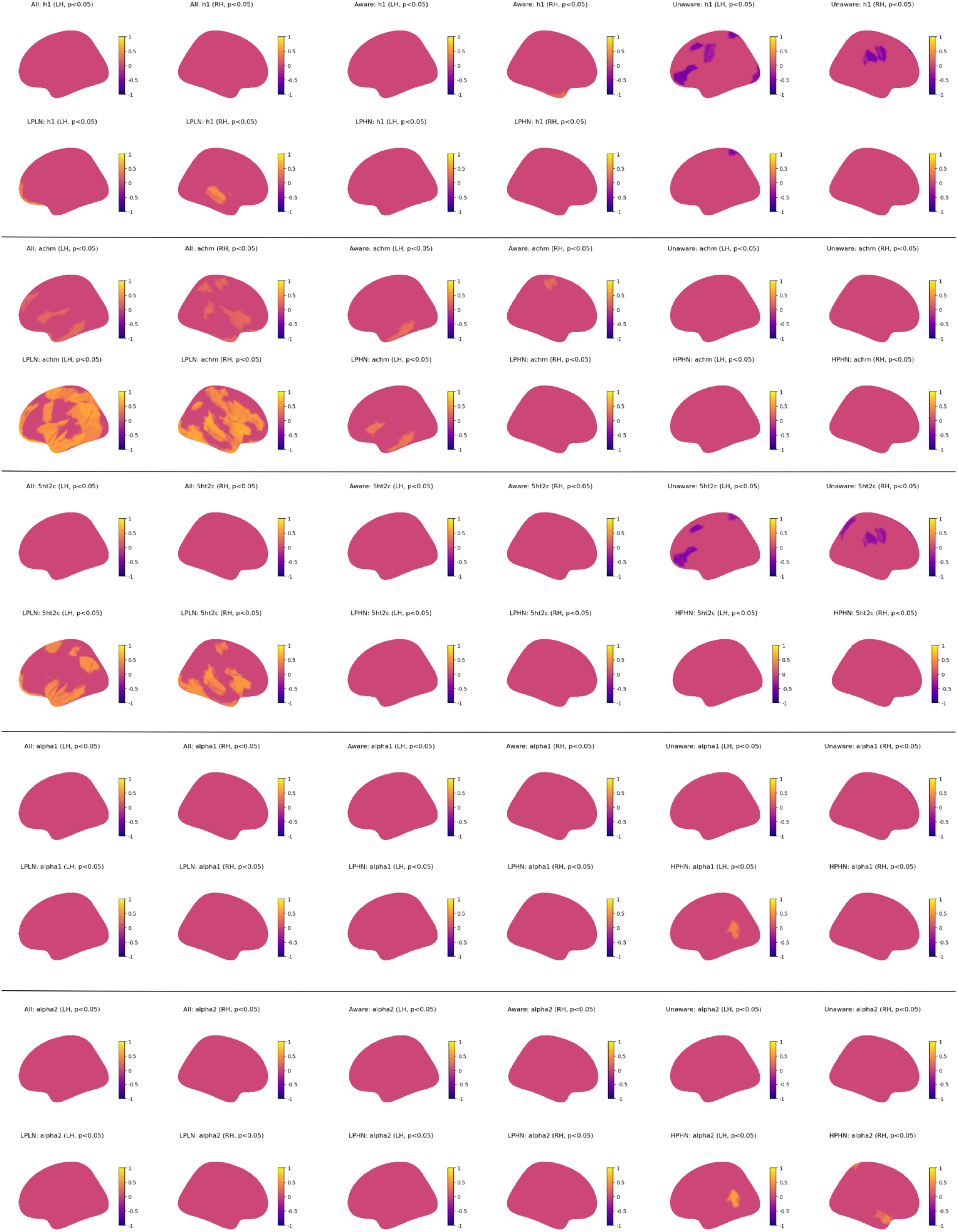
Significant correlations of regional fluidity and receptor occupancy. Orange regions indicate positive correlations and purple indicating negative correlations of the side-effects related receptors.

**Fig. A7.**
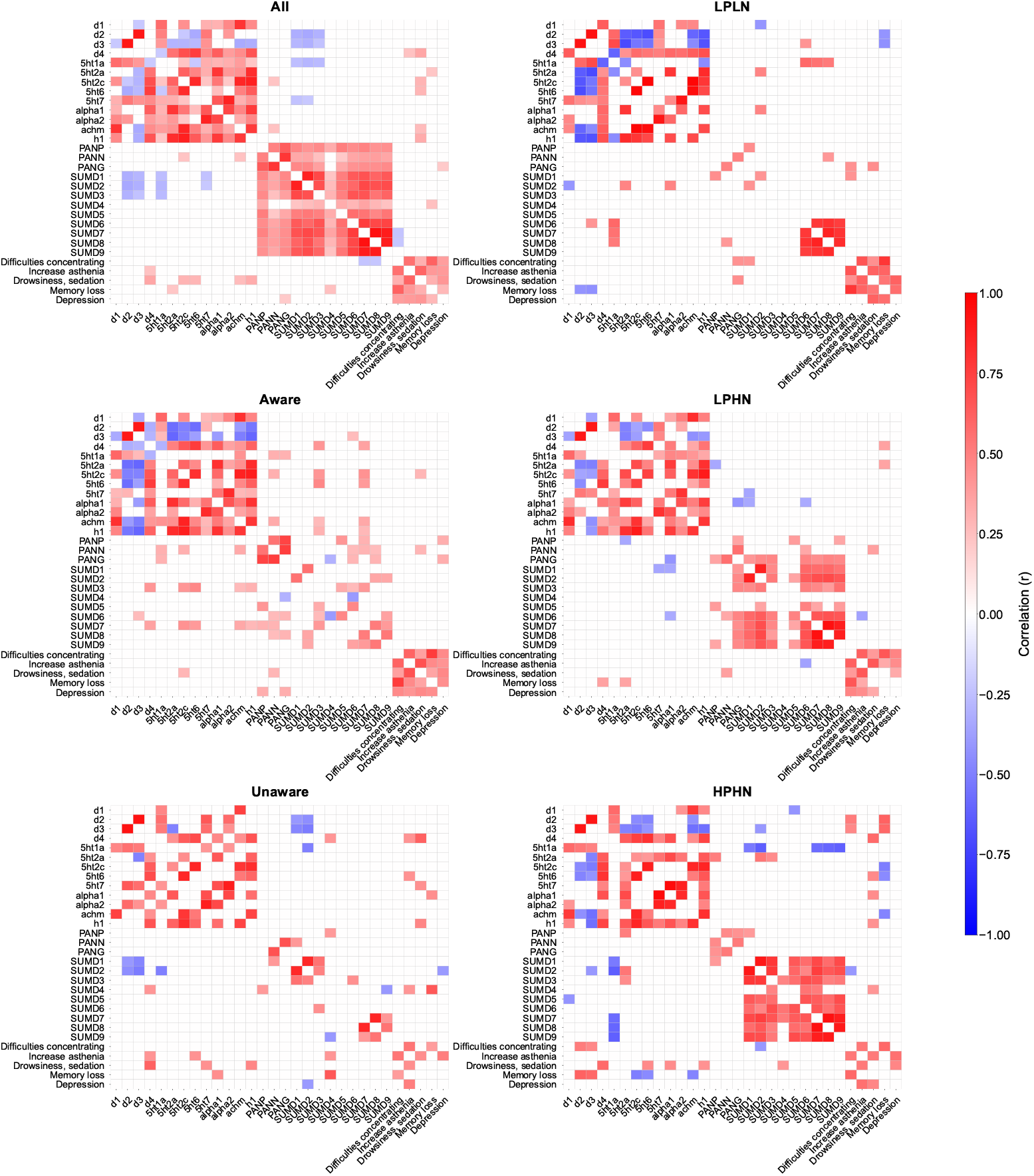
Intra-subtype significant correlations of receptor occupancy and clinical variables (Pearson’s r, *p* < 0.05). Different groups exhibit different corelational patterns, portraying the direct relationship of clinical outcome and pharmacological treatment.

**Fig. A8.**
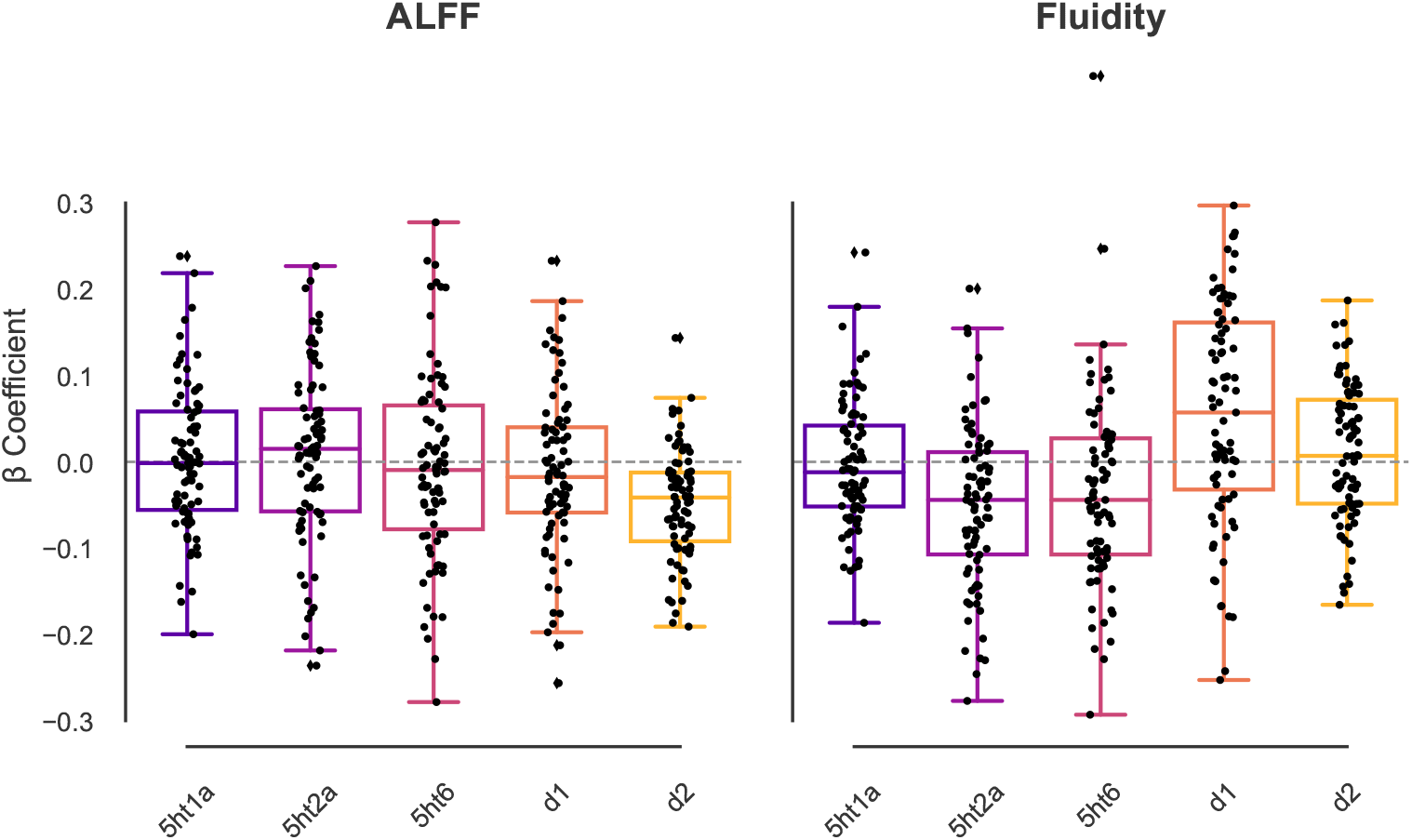
PLS Betta coefficient i.e spatial coupling of molecular architecture and regional features, fluidity, and ALFF in the whole population.

**Fig. A9.**
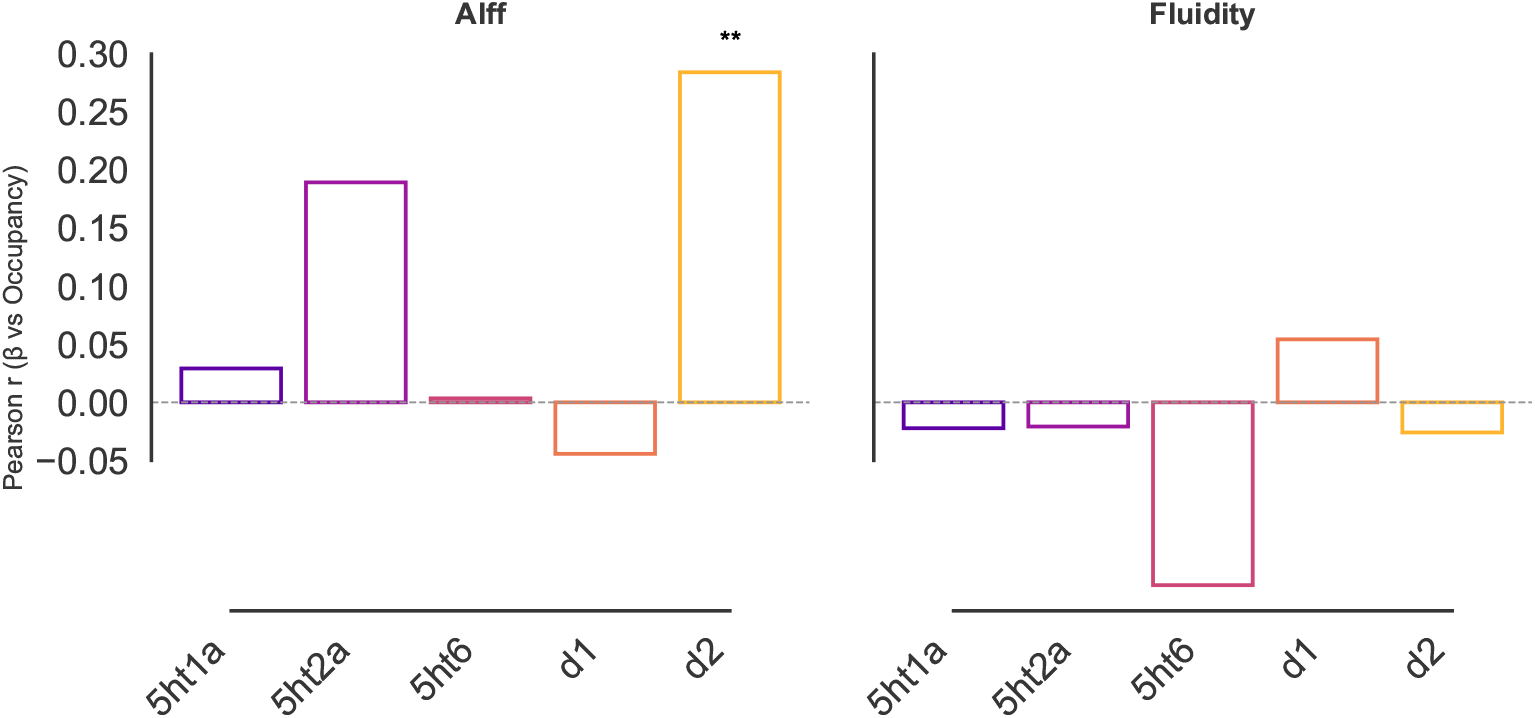
Correlations between molecular-functional coupling and receptor occupancy at in whole population level. Significant correlations (*p* < 0.05) in the D_2_ receptor occupancy and molecular-functional coupling, pointing to the more therapeutically targeted it is, the more functionally involved in regard to its molecular architecture in schizophrenia patients.

**Fig. A10.**
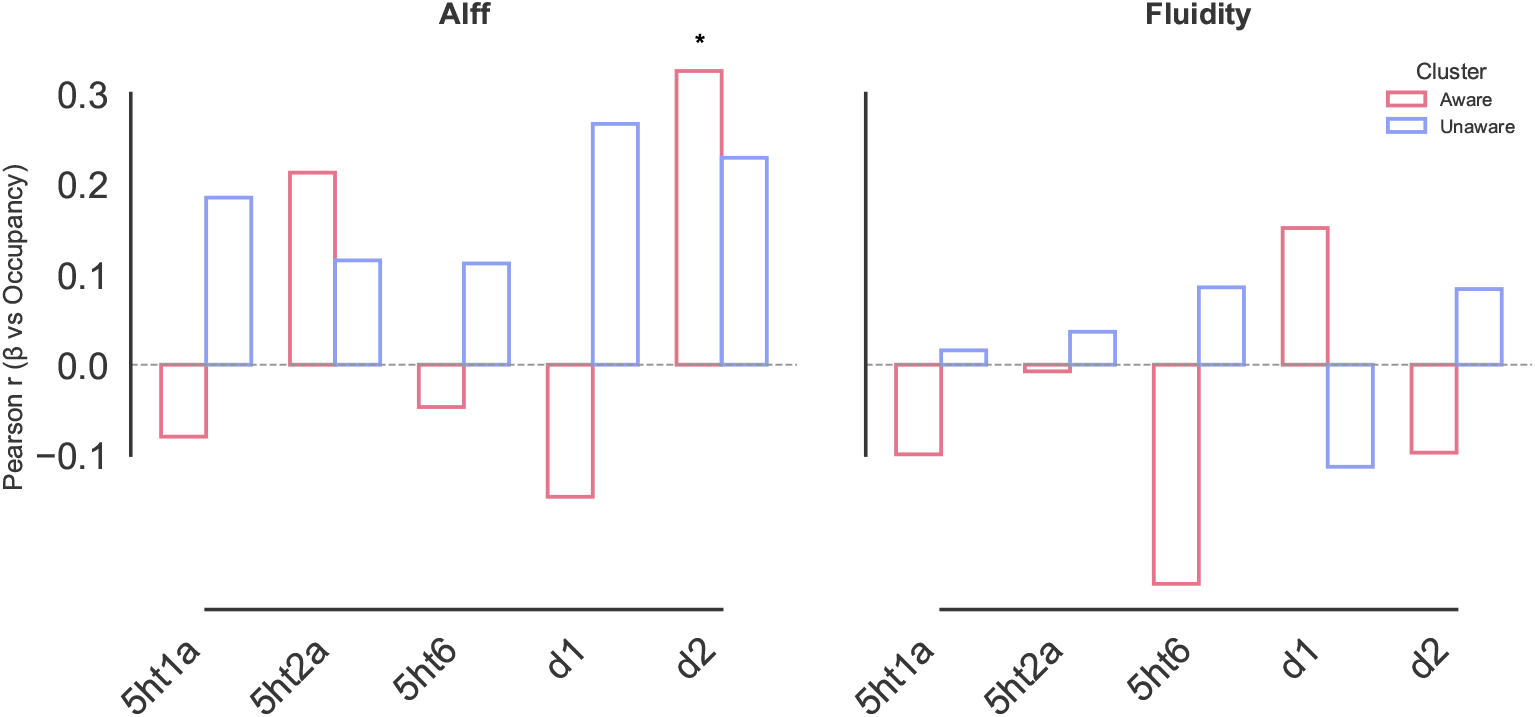
Within clinical data-driven subtypes, correlations between molecular-functional coupling and receptor occupancy. Significant difference in correlations (Man-Whitney U test FDR *p* < 0.05) in the D_2_ receptor occupancy and molecular-functional coupling emerged in the High Insight subtype. Addressing Fig A3, the more therapeutically targeted it is, the more functionally involved in regard to its molecular architecture contributed by the High Insight subtype.

**Fig. A11.**
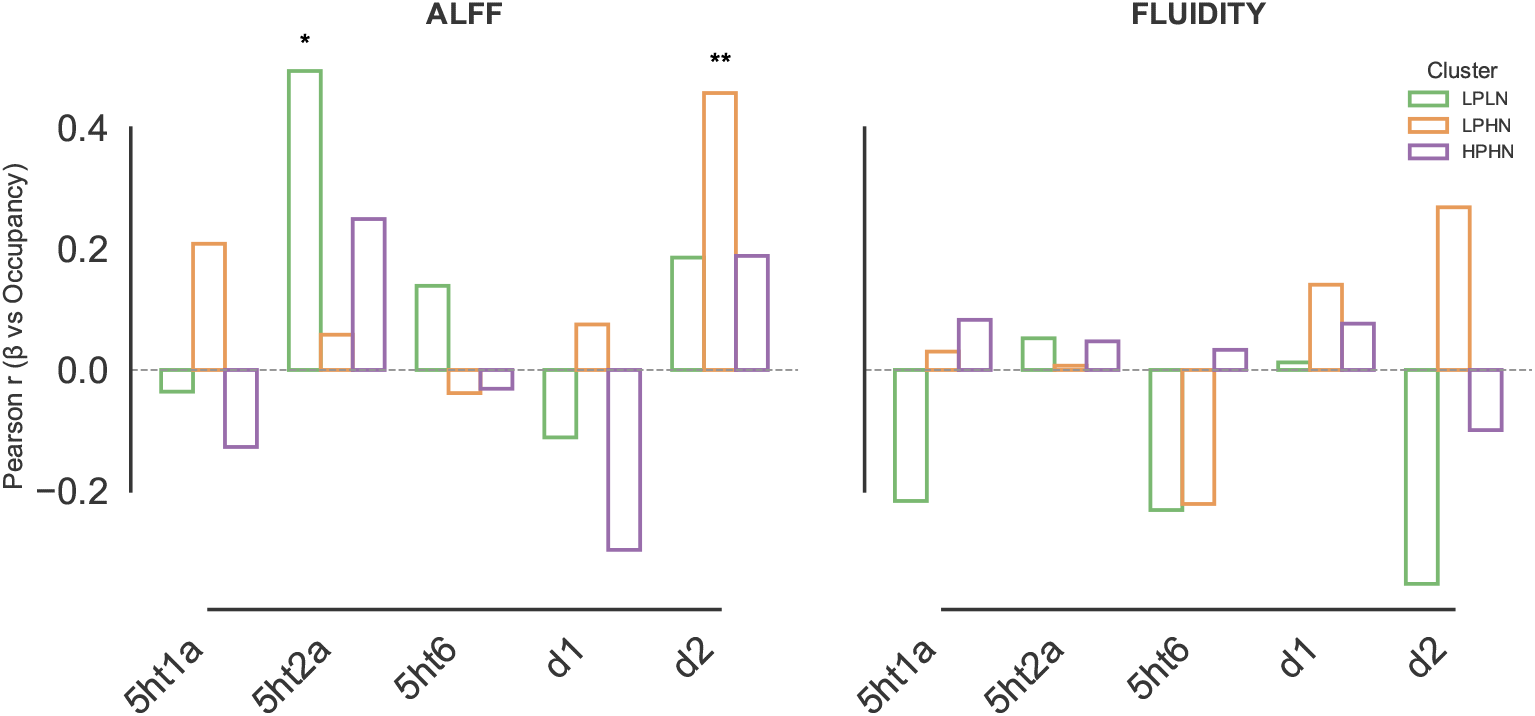
Within PANSS subtypes correlations between molecular-functional coupling and receptor occupancy. Significant correlations (*p* < 0.05) emerged in the D_2_ receptor occupancy and molecular-functional coupling in the LPHN subtype, showing that D_2_ is therapeutically targeted and functionally involved in the presence of negative symptoms, whereas 5HT_2_*a*’s involvement is significant in the LPLN subtype, suggesting that functional involvement or therapeutic neuromodulation is portrayed in subtypes with lower symptoms.

